# Landscape of Clinical Trials in Cancer Cachexia: Assessment of Trends From 1995-2024

**DOI:** 10.1101/2025.03.14.25323917

**Authors:** Ana Regina Cabrera, Kaitlyn Parker, Deena B. Snoke, Bart Hammig, Nicholas P. Greene

## Abstract

**Background:** Cancer cachexia, a multifactorial syndrome characterized by unintentional weight loss, is a frequent complication of cancer that impacts patients’ quality of life and survival.

**Methods:** In this retrospective review, we evaluated the landscape of clinical trials registered on ClinicalTrials.gov for the consideration of potential factors contributing to biological human heterogeneity in their design and analyses.

**Results:** Among clinical trials registered from 1995- 2024, we observed increased inclusion of female participants, but lack of reporting of sex as a biological variable. The majority (∼93%) of participants were of Caucasian descent. There was a substantial divergence in the diagnostic criteria and a wide range of tools employed to measure cancer cachexia. Lastly, few studies considered cancer type and stage as clinical variables.

**Conclusion:** Overall, a substantial gap remains in our knowledge of cancer cachexia in non-white individuals and in females. Ultimately, these underreported data across cancer cachexia clinical trials complicate the comparison and interpretation of clinical trials results, both in the broader human population and in specific cancer types. The current evolution of knowledge and new methodologies used for cancer cachexia assessment reinforce the need for a constant revision of the consensus definition and diagnosis criteria to align with current advances in our understanding of human heterogeneity in cancer cachexia.

## Background

Cancer is a critical global public health concern, with 9.7 million cancer-related deaths recorded worldwide in 2022 (1). Among all cancer types, lung, gastrointestinal, and pancreatic cancers are together estimated to comprise 38% of all cancer-related deaths in the United States (2). Cancer cachexia (CC), defined by Fearon et al. in 2011 as a multifactorial syndrome characterized by unintentional ongoing muscle loss which may occur with or without fat loss that is irreversible by conventional nutritional support (3), is a frequent complication of cancer which occurs at high rates in the cancer types mentioned above. CC is commonly clinically identified by decreased body weight or changes in body composition, which inevitably leads to impaired physical function. Because reliable body composition data are challenging, expensive, and impossible to retrospectively collect in a clinical setting, diagnostic criteria for CC have focused on body weight loss, with patients losing more than 5% of body weight or underweight patients (BMI<20 kg/m^2^) losing more than 2% of body weight in the past six months meeting criteria for CC (3). Clinical presentation of CC is a valuable metric to oncologists, as patients experiencing CC have increased fatigue (4), muscle weakness (5), treatment-related toxicities (6, 7), and decreased survival (5). Additionally, the frequency of CC is clinically significant, as its incidence increases in late-stage cancers (8), can be experienced in up to 80% of patients depending on cancer type (3, 9, 10) and is the main cause of mortality in ∼20-40% of cancer-related deaths (11).

Despite its prevalence and impact, efforts to develop preventative and therapeutic treatments for CC, including dietary supplements, exercise, and pharmaceutical drugs have had minimal success to date. The scientific community has acknowledged that these challenges may be due to the widespread use of preclinical models that may not faithfully recapitulate the human condition (12, 13), especially when extrapolated across cancers and other biological variables. Additionally, characteristics of CC can vary significantly depending on cancer type, tumor location, metabolism, mutational identity, and stage (7), which, due to the necessity of repeatability in disease models, is often not well reflected in preclinical studies. This is further complicated by factors intrinsic to human heterogeneity, which leads to significant variation in CC symptoms among individuals. One such factor that has gained traction in recent years is the effect of biological sex on CC characteristics (14). Clinically, males appear to exhibit well-described signs of CC earlier during cancer progression and undergo more significant declines in muscle mass in comparison to females (14). The observation that males adhere to this more ‘typical’ pattern of CC is partially due to the fact that historically, the majority of preclinical studies have utilized male mice or do not address biological sex as an experimental variable, and thus our understanding of the manifestation of CC in females is limited (15). Additionally, it is currently not feasible to capture the complexities of human genetic, environmental, and social factors such as race and ethnicity in preclinical models. However, recent patient data suggest that race and ethnicity demographics do impact disease outcomes such as CC incidence and mortality (16–19), underscoring the importance of studying diverse populations in clinical studies and trials to understand the genetic and environmental factors, including health disparities, contributing to disease outcomes. Additionally, the breadth of diagnostic measures reported across studies in human patients may contribute to inconsistencies in the literature that prevent pooled or comparative analyses across trials. The aim of the present review was to conduct a comprehensive assessment of CC clinical trials (CTx) between 1995-2024 and evaluate how these studies consider potential factors, such as demographic characteristics, intervention type, cancer type, and reported measures, contributing to heterogeneity of patient populations in their design, analyses, and interpretation. In doing so, we identify effective measures used by these studies and areas where alternative methods and considerations should be employed to address gaps in knowledge. Ultimately, the goal of this assessment is to aid in the advancement of future research initiatives by the design of more comprehensive and effective CTx in CC.

## Methods

This is a retrospective systematic review to analyze CTx investigating CC registered on ClinicalTrials.gov.

### Eligibility Criteria

Inclusion criteria were interventional and observational CTx investigating CC in child and adult subjects, both male and female, published on ClinicalTrials.gov. Trials were included if CC, cancer-induced sarcopenia, or cancer-induced muscle wasting were the primary reason for the study. Completed trials, active trials, and terminated trials were included. Exclusion criteria included trials that were unrelated to CC and trials measuring non-cancer induced cachexia. No trials were excluded based on country, region, total enrollment, trial start or completion date.

### Information Sources

The primary data source for this research was ClinicalTrials.gov, a comprehensive database providing information about clinical research studies conducted throughout the world. For trials listed without study results, PubMed was utilized to search for published manuscripts including results associated with them.

### Search Strategy

This search was conducted through ClinicalTrials.gov from December 2023 to August 2024, regardless of country and focusing on the condition/disease filter in the search generator. Terms related to CC, including “cancer cachexia”, “cachexia”, “sarcopenia”, and “muscle wasting” were searched one at a time. If “cachexia”, “sarcopenia”, or “muscle wasting” was the primary search term in the condition filter, “cancer” was added as a secondary search term in the other terms filter. No terms were added to the intervention/treatment filter or location filter. All boxes under study status were selected. The “all” option was selected under the sex filter. All age ranges were selected under the age filter. “Early Phase 1”, “Phase 1”, “Phase 2”, “Phase 3”, and “Phase 4” were selected under the study phase filter. Both “Interventional” and “Observational” were selected under the study type filter. “With results” and “Without results” were selected under the study results filter. No options were selected under the study documents, funder type, and date range filters. No additional information was input into the “More ways to search” section. When utilizing PubMed, the CTx official name followed by the study director’s name were searched.

### Data Collection Process

Data from eligible CTx were extracted from ClinicalTrials.gov. Official study title, ClinicalTrials.gov ID (NCT number), sponsor, study director, study start date, actual or estimated study completion date, study status, study type, enrollment, intervention(s), participant demographics, primary and secondary outcome measures, the type(s) of cancer included, CC definition used, and any listed publications were collected as reported on ClinicalTrials.gov. The same data were extracted from trials that lacked results or other data on ClinicalTrials.gov but were found in published manuscripts on PubMed. Published manuscripts were matched to CTx on ClinicalTrials.gov by NCT number. Principal investigators and participation enrollment were used as secondary confirmation to ensure the manuscript discussed the clinical trial of interest.

### Data Synthesis

The extracted data was grouped based on a variety of measures, including start and completion date of the trial, the treatment/intervention, type of cancer(s), CC definition used, and outcome measures. The list of CTx included is in **Supplemental File 1**.

### Figures and Tables

Figures were made with Biorender or R (v4.4.1). Tables and database were managed with Microsoft365 Word and Excel.

## Results

The searches conducted in ClinicalTrials.gov generated 209 trials using “cancer cachexia”; “cachexia” + “cancer” generated 226 results; “sarcopenia” + “cancer” generated 126 results; and “muscle wasting” + cancer” generated 143 results. After removing duplicates, 491 individual trials remained. After title and abstract screening, 300 trials did not meet the inclusion criteria. Therefore, the remaining 191 trials were included in this analysis. Of these, 106 trials had the status of complete, 10 of which had results published on ClinicalTrials.gov and 42 had published results in manuscripts available from PubMed, resulting in 52 completed trials with results. The remaining 54 completed trials did not have results identified on either ClinicalTrials.gov or PubMed. In addition, 62 trials were active, and 23 were terminated or withdrawn before completion at the conclusion of the search (**Supplemental File 2**). The reasons for termination are listed in **Supplemental File 3**, with none of the trials being terminated by safety boards. Moreover, most of the CTx (116, 60.7%) did not have a phase stated, 15 (7.9%) were in phase 1, 31 (16.2%) were in phase 2, 20 (10.5%) were in phase 3, 2 (1.0%) in phase 4, and 7 (3.7%) were in between two different phases (**Supplemental File 4**).

### Clinical Trial Demographics

We generated an overview of demographic composition of patients and how it was reported based on the 52 CTx with published findings. A summary of the reported demographics for age, biological sex, race, and ethnicity is compiled in **Figure 1a**.

**Figure 1.**
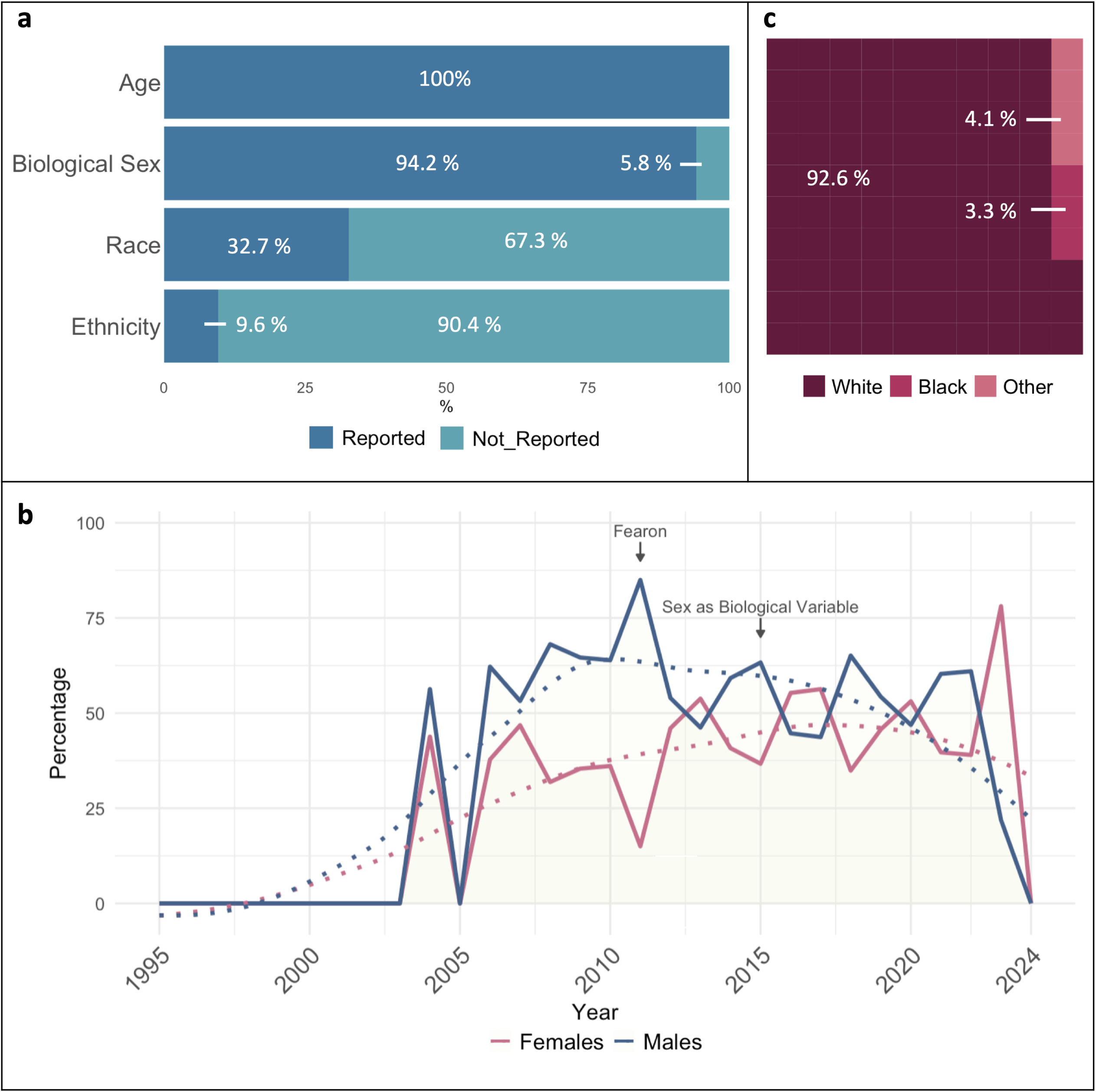
Demographics reported in clinical trials completed with results published from 2004-2023. **a)** Percentage of clinical trials including demographic data for age, biological sex, race, and ethnicity in the study population. **b)** Percentage of females and males included in clinical trials with reported results across years by clinical trial completion date (2004-2023). **c)** Percentage of White, Black, and Other races reported; others include 0.09% Native Hawaiian/Pacific Islander, 0.52% Asian, 0.91% Pardo, and 2.56% unknown (data not shown). “Fearon” marker indicates the year of the publication of the cancer cachexia consensus definition. “Sex as Biological Variable” indicates the year of the NIH expectation to consider sex as a biological variable in NIH-supported preclinical and clinical studies.

From the 52 CTx with results reported, 100% reported the ages of participants; of those, 50 trials (96.2%) reported the mean or median age of patients, ranging from 11 to 75 years old. From these 52 CTx, eight trials (15.4%) reported patient numbers by age categories (<18, 18-64, and >64). A total of 49 trials (94.2%) reported biological sex of participants and only one from the CC-CTx completed with results (1.9%) reported clinical outcomes by biological sex (NCT02148159). Among all trials with sex reporting, there were 3,103 females (40.4%) and 4,573 males (59.6%). In addition, we calculated the percentage of female and male participants within each study and the mean percentage had a similar distribution with 43.5% females and 56.5% males. However, we observed different trends across years. We noted an average of 38.7% females and 61.3% males in CTx with results reported from 2004 to 2014, while the average reported in CTx from 2015 to 2023 showed average distributions closed to equality with 48.8% of female and 51.2% of male participants (**Figure 1b**). In addition, 17 trials (32.7%) reported race of participants. Of these trials, all (100%) reported the participation of White patients. Fourteen trials (82.4%) reported the participation of Black or African American patients, six trials (35.3%) included Asian patients, two (11.8%) reported participation of Native Hawaiian/Pacific Islander individuals, one (5.9%) included Pardo patients, 9 (52.9%) included an unknown or mixed category, and no trials included American Indian/Alaska Native patients. Among all trials reporting race, there were 2,135 (92.6%) White patients, 77 (3.3%) Black or African American patients, 21 (0.9%) Pardo, 12 (0.5%) Asian patients, 2 (0.1%) Native Hawaiian/Pacific Islander patients, and 0 Native American/Alaskan Native patients or other races reported (**Figure 1c**). No trials reported clinical outcomes by race in a subgroup analysis. In addition to race, only five trials (9.6%) reported the ethnicity of participants. Among trials reporting ethnicity, 100% reported Hispanic/Latino patients making a 0.7% of the total population included in the CTx completed with results. Besides Hispanic/Latino participants, no other specific ethnicities were reported. No trials reported clinical outcomes by ethnicity in a subgroup analysis. Overall, CTx include both sexes but fail to report sex as a biological variable. Additionally, there is a significant knowledge gap in the study of CC among races and ethnicities other than the White population.

### Cancer Cachexia Definition and Classification

Despite the existence of a consensus definition of cachexia, the application of the diagnostic criteria remains only partially implemented in clinical practice. Therefore, we decided to classify the CTx according to the diagnostic criteria used to define CC. Our analysis demonstrated that of all trials included in this study (n = 191), 61.8% defined how they classified CC (**Figure 2a**). A significant portion of the studies (64, representing 33.5%) used the Fearon et al. consensus definition (3), stating a degree of loss and time frame for its occurrence. Studies that used weight loss greater than 5% in the past six months as their criteria, or weight loss greater than 2% in individuals already showing depletion with body mass index (BMI) <20 kg/m^2^ or skeletal muscle mass loss were classified as ‘Fearon’. Studies that defined CC in a different way (54, 28.3%), were classified as ‘Other’. Other criteria included different consensus definitions such as the Asian Working Group for Sarcopenia (AWGS) (20) European Society for Clinical Nutrition and Metabolism (ESPEN) (21), or European Working Group on Sarcopenia in Older People (EWGSOP2) (22). Other definitions included variations in time statements (1-12 months) or no statement regarding time frame, different body weight loss degrees (from 2-20%) or no statement about loss degree, other BMI ranges, or definitions by caloric intake or anorexia. Some other research groups utilized scores and tools such as the Edmonton Symptom Assessment Score (ESAS), Malnutrition Universal Screening Tool (MUST), Eastern Cooperative Oncology Group (ECOG), Karnofsky Performance Index, among others. Lastly, 74 (38.2%) CTx did not specify how they accounted for CC in their study. The distribution of the CC definitions used across years (1995–2024) is represented in **Figure 2b**. Although the Fearon consensus definition was published in 2011, the involuntary weight loss of ≥ 5% within the previous six months has been employed as a CTx criterion in studies initiated as early as 2005 (NCT00219817, NCT00267358, NCT00899158) (23). It is noteworthy that despite having several criteria and a consensus definition to diagnose CC, numerous studies fail to describe the diagnostic criteria used.

**Figure 2.**
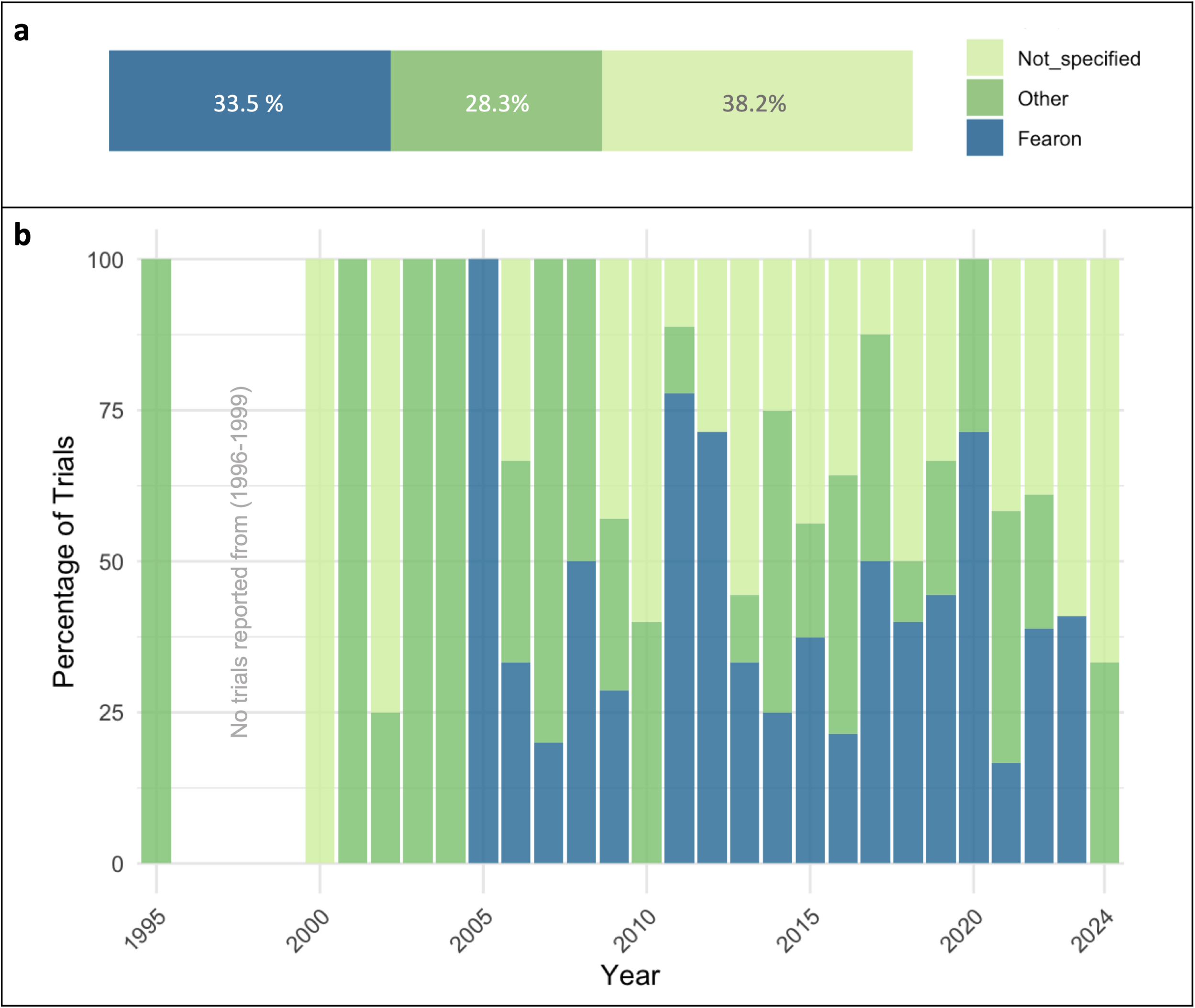
Cancer cachexia diagnostic criteria used in trials from 1995-2024. **a)** Studies that used weight loss greater than 5% in less than six months as their criteria, or weight loss greater than 2% in individuals already showing depletion with body mass index (BMI) <20 kg/m^2^ or skeletal muscle mass loss were classified as ‘Fearon’; studies defining cachexia in a different way were classified as ‘Other’. **b)** Distribution of the cachexia definitions used across years.

The current status of CTx stratified by their CC definition criteria and terms used for trial classification are summarized in **Supplemental Files 5 and 6**. Of 52 CTx completed with results, 29 (55.8%) specified the use of the Fearon et al. consensus definition. Fourteen trials (26.9%) used definitions other than the Fearon et al., with six of those trials beginning after the consensus definition was established in 2011. Nine trials (17.3%) did not specify the criteria used for how they defined CC. Moreover, five trials (9.6%) were classified under “sarcopenia”, nine trials (17.3%) were classified under “weight loss”, four trials (7.7%) were classified under “muscle wasting” or “muscle loss”, and four trials (7.7%) were classified under “malnutrition” or “anorexia”. Within the completed trials without results, nine trials (16.6%) specified the use of the 2011 Fearon et al. consensus definition. Nineteen trials (35.2%) used definitions other than the Fearon et al. definition, with 11 of those trials beginning after the consensus definition was established in 2011. Twenty-six trials (48.2%) did not specify how they defined CC. From these trials without results, six trials (11.1%) were classified under “sarcopenia”, ten trials (18.5%) were classified under “weight loss”, six trials (11.1%) were classified under “muscle wasting” or “muscle loss”, and three trials (5.5%) were classified under “malnutrition” or “anorexia”. For the CTx that were still active at the completion of the search, 24 (38.7%) specified the use of Fearon definition and eight trials (12.9%) used other definitions. Thirty (48.4%) did not specify how they defined CC for their study. Among these active studies, 16 (25.8%) have been classified under “sarcopenia”, one trial (1.6%) under “weight loss”, nine trials (14.5%) under “muscle wasting” or “muscle loss”, and one trial (1.6%) classified under “malnutrition” or “anorexia”. From the withdrawn trials, two (8.7%) used Fearon definition, 13 (56.5%) used other definitions, and eight (34.8%) did not specify their criteria to classify CC. Three trials (13.0%) were classified under “weight loss”, one trial (4.4%) was classified under “muscle wasting” or “muscle loss”, and one trial (4.4%) was classified under “malnutrition” or “anorexia”.

### Sample Size, Cancer Types, and Clinical Approaches Across Clinical Trials

We characterized study participants among CTx to determine the number of registered participants and the cancer type and stage studied in each trial. Across all trials (n = 191), 72 (37.7%) enrolled between 1-50 patients, 49 trials (25.7%) enrolled between 51-100 patients, 51 (26.7%) enrolled between 101-500 patients, six (3.1%) enrolled between 501-1000 patients, four (2.1%) enrolled more than 1000 patients, and nine trials (4.7%) did not specify the total enrollment, most of which were from withdrawn trials. The distribution of patients’ enrollment per clinical trial status is shown in **Supplemental Files 7A and 7B**.

Analysis of the CTx by cancer type show that 50.3% of trials studied CC in a specific type of cancer, 8.9% studied two cancer types, 5.2% studied three cancer types, 11.5% studied four or greater cancer types, and 24.1% did not state the cancer type considered for their study (**Figure 3a**). Among these CTx, CC has been studied more predominantly in lung and gastrointestinal related cancers (pancreatic, gastric, colorectal), either individually or in combination with other cancer types. In addition, between 10-35% of the trials did not report a concurrent anti-cancer treatment despite that it is standard of care (data not shown), and the majority did not report the cancer stages represented in the study (146, representing 76.4%). Of those trials reporting patient cancer stage (n=45), 53.3% investigated CC in advanced stage cancers (stage III-IV), while other stages were reported between 2-15% (**Figure 3b**). **Supplemental File 8** details the stratification of cancer type and stage per CTx status.

**Figure 3.**
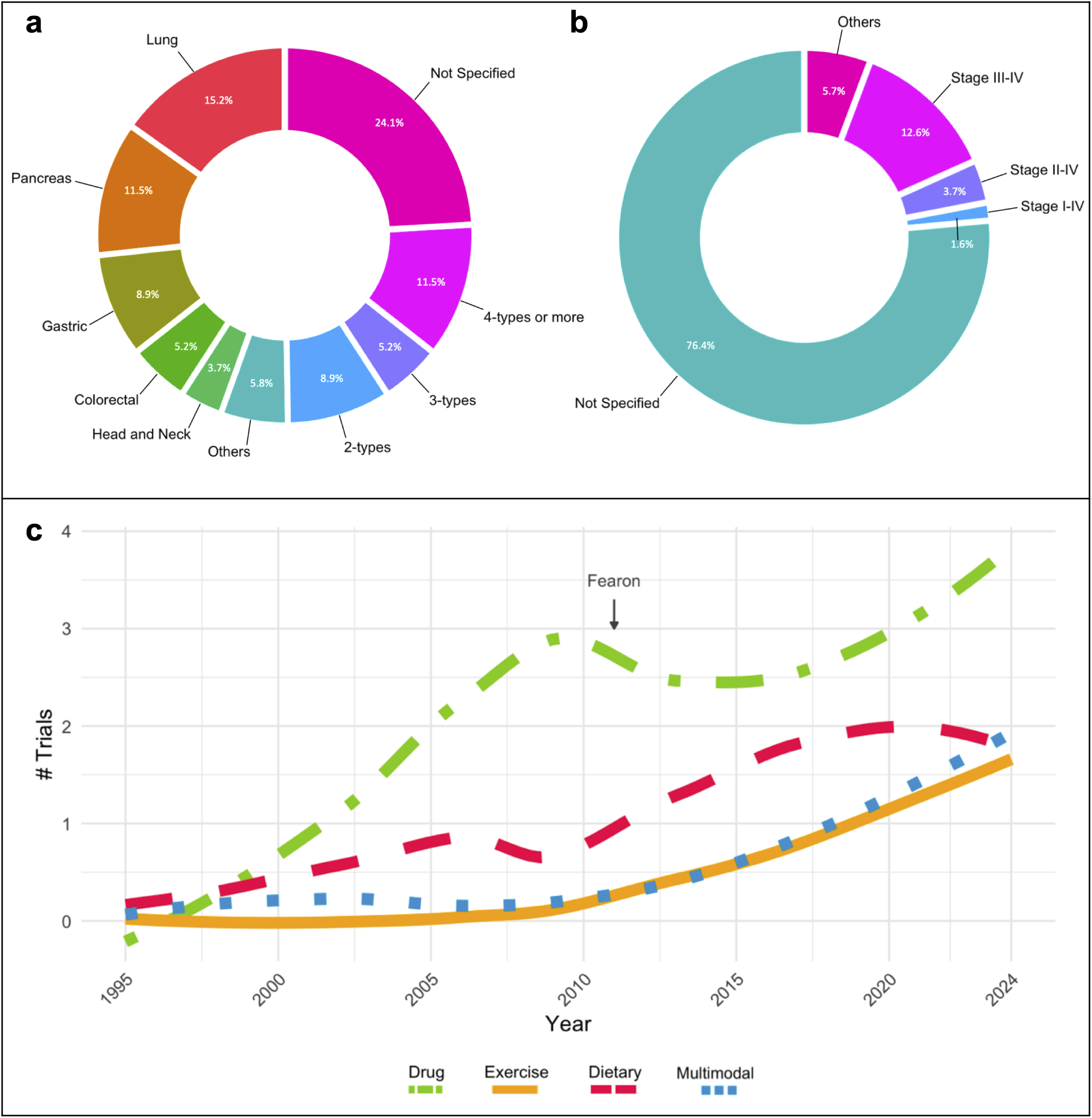
Study population and approach. **a)** Cancer type and **b)** stages reported in clinical trials related to cancer cachexia. **c)** Clinical trials approach. LOESS (locally weighted smoothing) regression analysis from pharmacological (drugs), exercise, dietary, and multimodal interventions across years (1995-2024) investigated in cancer cachexia clinical trials. “Fearon” marker indicates the year of the publication of the cancer cachexia consensus definition.

CTx studying the effects of CC employed either an observational (26.7%) or an interventional (73.3%) study design. The most common intervention was pharmaceutical (33.5%, “drug” on the Figure), followed by dietary supplements (14.7%), exercise (7.9%), and different multimodal approaches (9.4%). Multimodal interventions included different combinations of dietary supplements, nutrition, exercise, drugs, counseling, and psychological support. The interventions used by CTx status are detailed in **Supplemental File 9** and a list of the drugs and dietary supplements utilized as intervention are detailed in **Supplemental File 10**. The frequency of intervention types over time among CC-CTx can be visualized in Figure 3c. These trends indicate that drug treatment interventions remain a popular approach over time. Additionally, a noticeable increase in the use of exercise and dietary interventions is observed following the Fearon consensus definition in 2011.

### Outcome Measures and Tools Used

To understand how CTx assessed CC, we summarized the evaluated parameters and the tools that have been used to measure them. First, we looked for body weight, a parameter considered in most CC definitions, and only 42.4% of studies included body weight as an outcome measured, from these, only 20% were prior to Fearon’s CC definition (**Figure 4a**). A total of 62.3% of studies report the use of body composition across all trials (with only 18.5% of them prior Fearon’s CC definition in 2011), with 25.7% reporting muscle mass, 23.6% reporting lean body mass, and 11.0% reporting fat mass, among others (**Figure 4b**). To assess body composition, computed tomography (CT) was the most used technique (18.3%), followed by bioelectrical impedance analysis (BIA, representing 17.3%) and dual energy x-ray absorptiometry (DXA, 12.6%). Physical function was an outcome measure in 50.3% of CTx, including measures of muscle function or muscle strength (29.3%), physical activity levels (13.6%), and fatigue (7.3%) (**Figure 4c**). To assess physical function, 26.2% of the trials used hand grip strength and 13.1% used a 6- or 10-minute walk test, with other methodologies used less frequently. Quality of life (QoL) was specified by 37.2% of the trials (**Figure 4d**), and it was mostly evaluated by the ECOG Performance Status Scale (33.0%), the European Organization for the Research and Treatment of Cancer QoL Questionnaire (EORTC QLQ, 20.9%), and by the Functional Assessment of Anorexia/Cachexia Treatment (FAACT, 13.6%), among others. Nutrition and appetite were mentioned by 30.4% of the CTx and dietary recall or food diaries were the methodologies most used (9.4%) (**Figure 4e**). Molecular measures were mentioned in 45.5% of the studies, with ∼45% of them stated in CTx studying CC in the last 5 years (**Figure 4f**). Of those studies reporting molecular measures, most studies assessed blood samples (44.5%) while some reported use of muscle biopsies (11.0%), the specific targets measured are listed in **Supplemental File 11**. Lastly, 21.5% of CTx specified the measure of safety and tolerability, and 19.4% reported survival (**Figures 4g and 4h**). Outcomes measured and other tools that have been used to a lesser extent are detailed by clinical trial status on the **Supplemental Files 12 and 13**.

**Figure 4.**
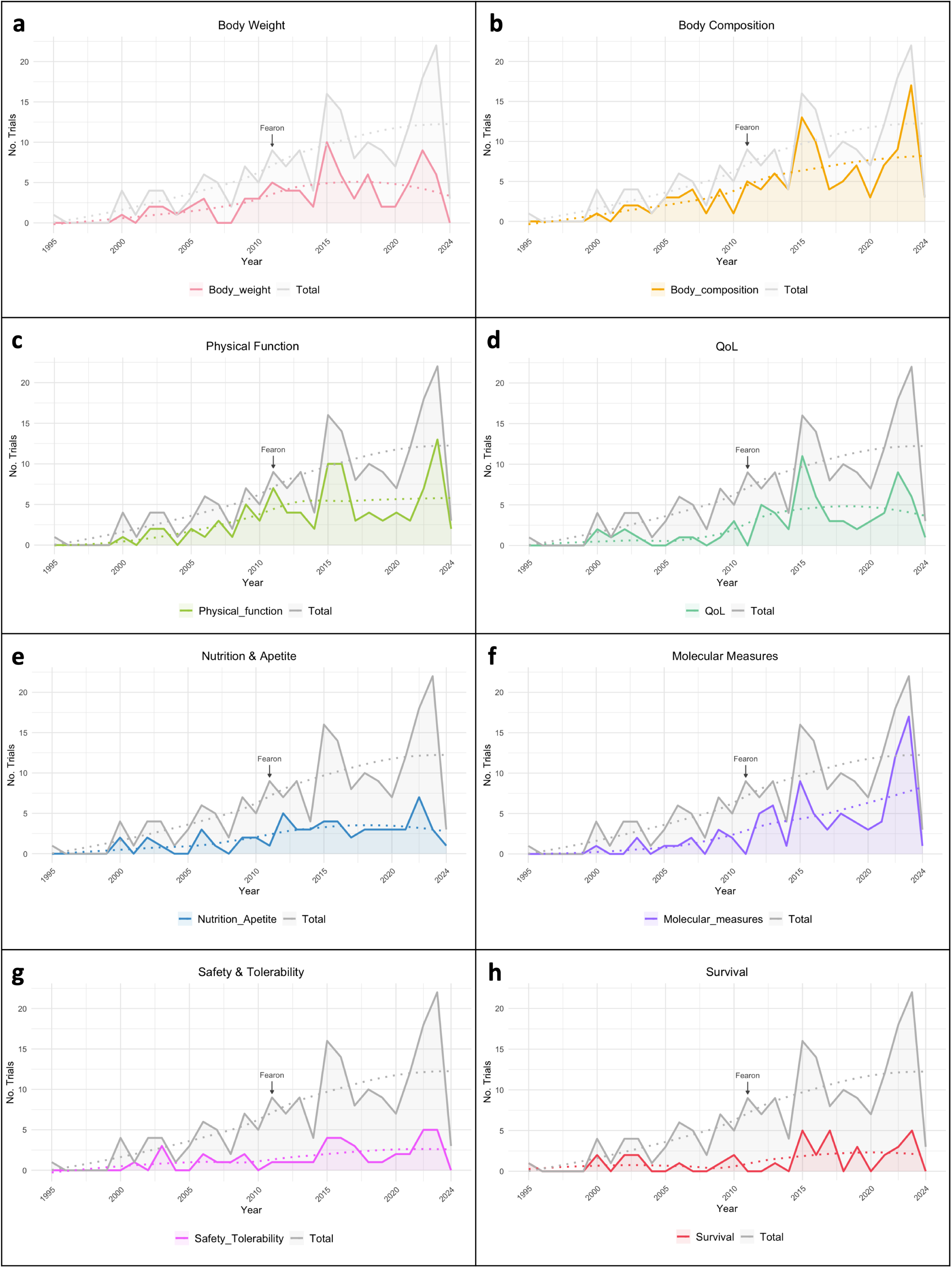
Clinical trials outcomes across years (1995-2024). Number of trials and LOESS (locally weighted smoothing) regression analysis from **a)** body weight, **b)** body composition, **c)** physical function, **d)** quality of life (QoL), **e)** nutrition and appetite, **f)** molecular measures, **g)** safety and tolerability, and **h)** survival measured in cancer cachexia clinical trials. “Fearon” marker indicates the year of the publication of the cancer cachexia consensus definition.

## Discussion

While significant progress has been made in the understanding of the pathogenesis of CC and in establishing potential therapeutic interventions, CC significantly decreases quality of life and remains a main cause of death for many cancer patients, while successful treatment strategies have not yet been identified. Through evaluation of CC CTx, we assessed aspects of human heterogeneity that are not well-addressed in carefully controlled animal studies and assess changes in study approaches and improvements in these metrics over time. Our findings suggest a large knowledge gap in how CC differs across patient populations and how this impacts health outcomes. Expanding CTx to improve representation across patient populations may improve progress toward effective treatments for CC.

### Reporting of Demographic Factors

It was not until 1993 that in the United States (U.S.), the National Institutes of Health (NIH) Revitalization Act was passed requiring recruitment and inclusion of women and racial and ethnic minorities in clinical research studies. In 2011, almost 20 years afterwards, the Institute of Medicine of the National Academies suggested reporting results by biological sex be required for the publication of CTx results, as lack of reporting had significantly slowed the advancement of women’s health (24, 25). Recognizing sex-based biological differences is essential for understanding mechanistic and pathophysiological divergence during the onset and progression of pre-clinical studies of CC (14, 15). Therefore, the acknowledgment of women’s inclusion in CTx alone is insufficient. In 2015, the NIH announced the requirement for sex to be considered as a biological variable in NIH-supported preclinical and clinical studies. Herein, we identified that across all CC-CTx completed with results, 5.8% did not report sex at all, while 59.6% of the total patient population was male. This is consistent with recent findings that across studies assessing longitudinal body composition in patients with lung cancer, 65% of all study participants were male (26). Additionally, in the present review we find that between 2015-2024, the percentage of enrolled females increased to 48.4% of patients, reflecting increased equality of enrollment of males and females in CTx over time suggesting enhanced equity. At present, significant advancements have been made in understanding the pathogenesis of CC, but still very little is known about how biological sex as a variable affects patient outcomes with CC. Despite improved equality of enrollment, trends are yet to come to fruition for the consideration of biological sex as a variable in data analyses, as in twenty-nine years we only identify one CTx reporting sex subgroup analysis. Our findings highlight that despite many positive changes in the conduct of clinical studies, inequalities persist, likely contributing to suboptimal progress. Understanding sex as a biological variable in clinical CC will ultimately increase the ability to accurately model the complications of the human condition, and in turn, develop effective treatments improving health outcomes for both males and females with CC.

Approximately one-third of CTx completed with results reported race, while less than one-tenth reported ethnicity. Of CTx reporting race and ethnicity, there was a lack of racial and ethnic diversity among enrolled patients. Among CTx reporting race, 92.6% of all patients were White. In CTx reporting ethnicity, 100% focus solely on Hispanic/Latino individuals, neglecting other ethnicities. This homogeneity fails to reflect the true representation of the general population and highlights the scarcity of information about CC in non-white individuals, severely limiting the understanding of how race and ethnicity affect CC. Importantly, a recent study reports that Black patients were more frequently excluded from clinical studies due to comorbidities, rendering them ineligible for participation (27). Based on our findings, these eligibility criteria may exacerbate disparaged enrollment that expands beyond only PDAC patient populations and should be considered in future clinical studies. Through 2024, recent calls for applications from the National Cancer Institutes in the U.S. highlighted the dire need to understand cancer burden across population groups. Although contributions to this area of research are currently minimal, there have been significant recent findings that provide evidence for possible genetic, environmental, and socioeconomic differences across race and ethnicity warranting further studies. For instance, across all stages of lung cancer, Black race strongly increased the odds of experiencing pretreatment weight loss (17) and in patients with advanced lung cancer, Black race and Hispanic ethnicity presented with greater risk of developing CC (16). The Florida Pancreas Collaborative has made great strides in understanding racial and ethnic differences in pancreatic CC, reporting greater frequency of cachexia markers in Blacks with concurrent higher cancer incidence and mortality and shorter survival time (18, 19). Additionally, Hispanic/Latino patients experienced increased survival time compared to non-Hispanic White individuals and Black individuals (19). This suggests underlying genetic, environmental, socioeconomic, and/or yet unknown factors predisposing patients to increased risk of developing CC, which may differ beyond cancer type. Despite these observations, no CTx reported clinical outcomes by race or ethnicity in a subgroup analysis. As with biological sex, race and ethnicity should be included in subgroup analyses for clinical outcomes across CC studies; however, studies must be statistically powered for these analyses and thus recruitment of a racially and ethnically diverse study population is critical.

### Inconsistent Cancer Cachexia Definitions and Varied Diagnostic Criteria

Despite that a consensus CC definition was published in 2011 (3), we found considerable heterogeneity in the definition and diagnostic criteria used across CTx, with only 33.5% of studies using the consensus definition criteria to classify CC. Additionally, there have not been improvements in these metrics over time. Significant variations in classification persist, with a significant portion (38.2%) of CTx not reporting the details of their classification methods, which presents an enormous challenge in interpreting and comparing results across CTx. Additionally, a significant portion of studies use description of symptoms to define CC, including sarcopenia, muscle wasting/loss, anorexia, and malnutrition. The inconsistency in the terminology and diagnostic guidelines may lead to misdiagnoses, absent medical records, and inaccurate reports of cachexia incidence, likely concealing the visibility of the impact of CC on public health. Altogether, these gaps reflect a systemic oversight of clinically important characteristics of the syndrome. The lack of standardization demands action to unify criteria and reporting practices to enable meaningful data integration and cross-study comparisons that lead to cachexia research advancement.

### Consideration of Cancer Type Effects on Cancer Cachexia Patient Outcomes

We found that many CC-CTx have relatively small samples sizes, with most CTx in this review recruiting 100 participants or fewer. CTx on CC require many clinical assessments in patients who are often unwilling or unable to contribute to research, and thus it can be challenging to recruit patients on a large scale. However, efforts to recruit larger samples sizes could increase the ability to better understand CC in diverse populations and achieve statistical power required for subgroup analyses. Additionally, almost one-third of CTx encompassed in the present work included more than one cancer type in the study population. Recently, it has become more apparent that different cancer types likely exhibit different patterns of tissue wasting and progression. This is clear not only in clinical findings, but also in preclinical models of CC where different tumor types exhibit mechanistic differences during disease development and progression (15). Thus, combining different cancer types without offering stratified analyses likely conceals cancer type-specific changes. However, we report that 41.5% of completed CTx and 64.5% of active CTx focused on a single cancer type, suggesting the evolving knowledge of varying clinical cancer characteristics and treatments has informed the design of CTx to improve our understanding of cancer type-specific CC outcomes. In addition, although CC is commonly diagnosed at advanced cancer stages, 76.4% of CTx included in this review did not report a specific cancer stage. Moreover, 73.3% of the studies incorporated in this analysis were interventional. Consequently, the lack of consideration for cancer stage as a clinical variable may skew the results of the interventions, particularly by overlooking pre-cachectic states in which knowledge remains limited.

### Growing Focus on Exercise Interventions in Cancer Cachexia Clinical Trials

The use of pharmaceutical approaches to treat or prevent CC remains the most popular intervention among CC-CTx. Some of them report promising results, ameliorating some of the cachexia signs, such as Anamorelin and Ponsegromab (23, 28, 29). We observed a growing focus on exercise interventions in the study of CC, either alone or in combination with other treatment paradigms in recent years. As demonstrated by Schink et al. (NCT02293239) (30), a multimodal approach utilizing whole-body electromyostimulation (WB-EMS) combined with a light dynamic exercise training program alongside nutritional support, led to significant increases in skeletal muscle mass, body weight, and Karnofsky performance index over 12 weeks of training compared to the cancer control group (30). Highlighting exercise as a clinically relevant and promising strategy to mitigate the effects of CC. Additionally, there was a relative increase in the measure of physical function and quality of life, reinforcing the importance of maintaining muscle quality to enhance well-being in patients with CC. Along with a growing understanding of the molecular mechanisms of tissue wasting informed by preclinical studies, we observed an increase in the use of molecular measures in CTx. Broadly, we observed wide variation in the outcome measures reported, even those non-invasive, affecting the ability to analyze systematic reviews reinforcing clinical conclusions.

### Body Weight is Underutilized as an Outcome in Cancer Cachexia Assessment

Although body weight and composition are fundamental measures of CC, we identified a surprisingly low frequency of studies reporting body weight (42.4%) and body composition (62.3%). Indeed, body weight loss utility may be enhanced when considered alongside other metrics, as demonstrated by Martin and collaborators (31), a BMI-adjusted weight-loss grading system (grades 0 to 4) that accounts for median overall survival and unadjusted estimated hazard ratios (31). With more accurate tools for body composition measurement, such as CT and MRI, recent studies have brought attention to body composition changes across the cancer and treatment continuum within certain CC-associated cancer types (32, 33). Moreover, body composition, reported by CT, is often a primary outcome of CC-CTx, and sexual dimorphisms in the distribution of fat and muscle tissue in the thoracic and lumbar compartments must therefore be considered in the interpretation of body composition assessment. We found that active CTx are more frequently using these measurement tools, which will provide the field with a better understanding of body compositional changes during disease progression. In light of these findings, consistency of diagnostic criteria and outcome reporting across future CTx are essential in advancing our understanding of CC and improving clinical outcomes. To address this need, the Cancer Cachexia Society has convened a CTx Advisory Board, a multidisciplinary group of CC experts to provide transparent guidance on clinical trial design through a confidential and comprehensive review process.

## Conclusions

Overall, a substantial gap persists in our understanding of CC in non-white and female populations. The report of CC definition and transparency in diagnostic criteria, along with the absence of stratified variables such as cancer type and biological sex are deficient, complicating the comparison and interpretation of CTx results. Lastly, the arrival of new techniques and methodologies used for cachexia assessment reinforces the need for a regularly revised and updated consensus definition and diagnosis criteria to align with new research findings and cutting-edge technology.

## Limitations

In the findings we present in this review article it was challenging to determine the setting in which patients are recruited (e.g. oncology, palliative care) due to inconsistencies in reporting of patient population data and disease characteristics, which may influence the characteristics of the populations recruited for these studies.

Assessment of active, completed, and terminated CTx in this review reveals both the recent advancements and remaining gaps in clinical CC research. Addressing the lack of subgroup analyses based on biological sex is essential for comprehensive understanding of the differences in the development and progression of CC between males and females. Similarly, investigating the diverse manifestations of CC among various racial and ethnic groups is crucial for eliminating health disparities and developing treatment options that are effective in diverse populations. Standardizing CC diagnostic criteria across future clinical trials can enhance the reliability and effective comparability of trial findings. Additionally, small and heterogeneous study samples make drawing conclusions challenging, emphasizing a need for larger recruitment and modified trials designs that minimize the combination of cancer types, staging, and concurrent therapies within study populations. Current trends in intervention methods mirror advancing knowledge of CC, with corresponding changes in outcome measures. However, significant gaps and inconsistencies persist creating a dire need for continued improvement. By addressing these issues and proposing more effective and standardized study designs, we aim to contribute to ongoing advancement of our understanding of CC and ultimately improve patient outcomes.

## Data Availability

All data produced in the present work are contained in the manuscript and associated supplemental figures.

## Abbreviations

AWGS: Asian Working Group for Sarcopenia
BIA: Bioelectrical impedance analysis
BMI: Body mass index
CC: Cancer cachexia
CT: Computed tomography
CTx: Clinical trials
DXA: Dual energy x-ray absorptiometry
ECOG: Eastern Cooperative Oncology Group
EORTC QLQ: European Organization for the Research and Treatment of Cancer QoL Questionnaire
ESAS: Edmonton Symptom Assessment Score (ESAS)
ESPEN: European Society for Clinical Nutrition and Metabolism
EWGSOP2: European Working Group on Sarcopenia in Older People
FAACT: Functional Assessment of Anorexia/Cachexia Treatment
MUST: Malnutrition Universal Screening Tool
NIH: National Institutes of Health
QoL: Quality of life
WB-EMS: Whole-Body Electromyostimulation

## Ethics approval and consent to participate

Not applicable.

## Consent for publication

Not applicable.

## Availability of data and materials

Correspondence and requests for materials should be addressed to Nicholas P. Greene.

## Authors’ contributions

**ARC**: drafting all the figures and final manuscript, reviewing, and editing. **KP:** creating the database, drafting the manuscript, reviewing, and editing. **DBS:** creating figures, writing, reviewing, and editing. **BH**: conceptualization, reviewing, and editing. **NPG:** conceptualization, resources, supervision, reviewing and editing. All authors reviewed and approved the final manuscript.

## Acknowledgements

The authors thank all the faculty, staff, and students of the Exercise Science Research Center at the University of Arkansas.

## Supplementary Information

**Supplemental File 1.**
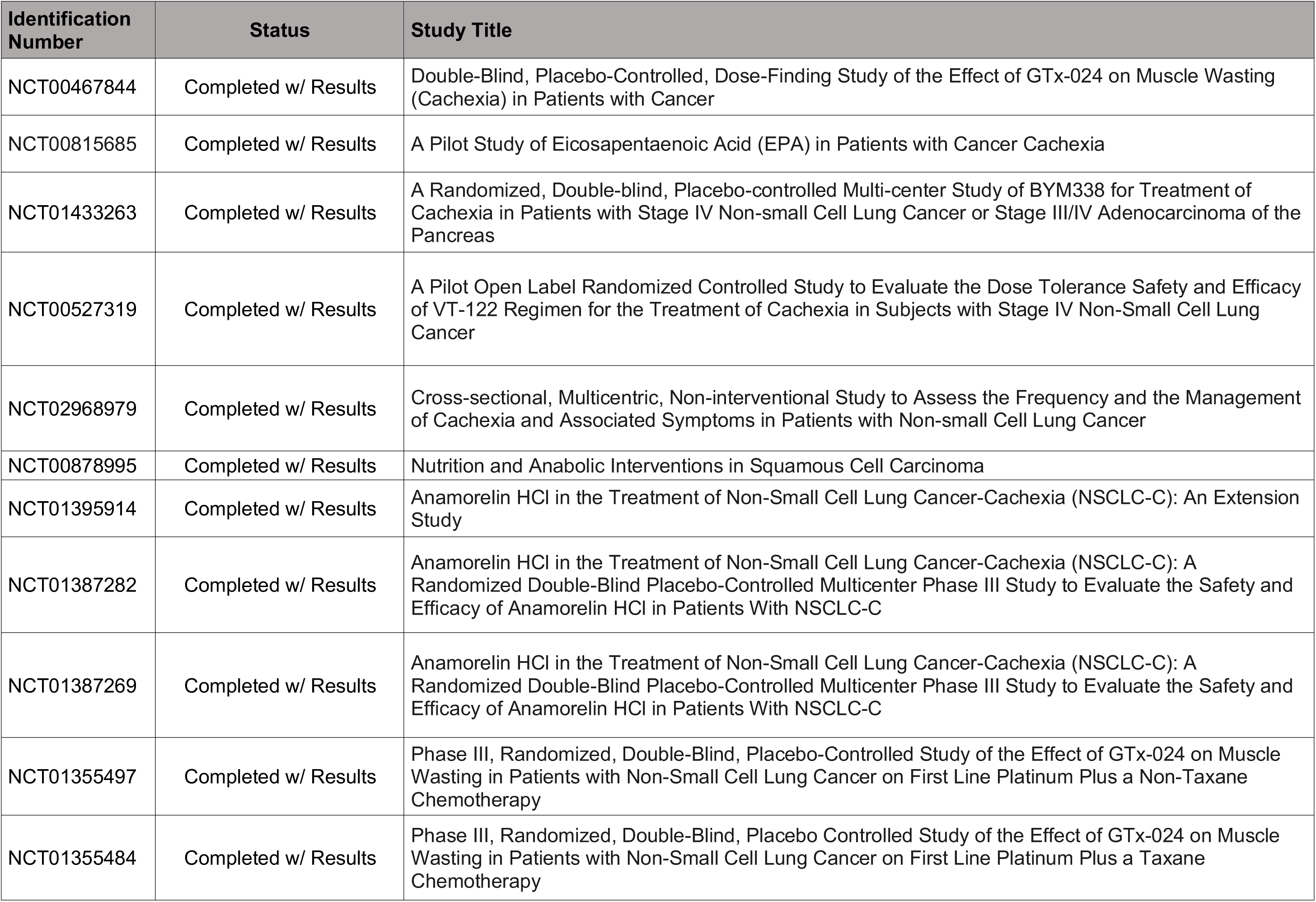
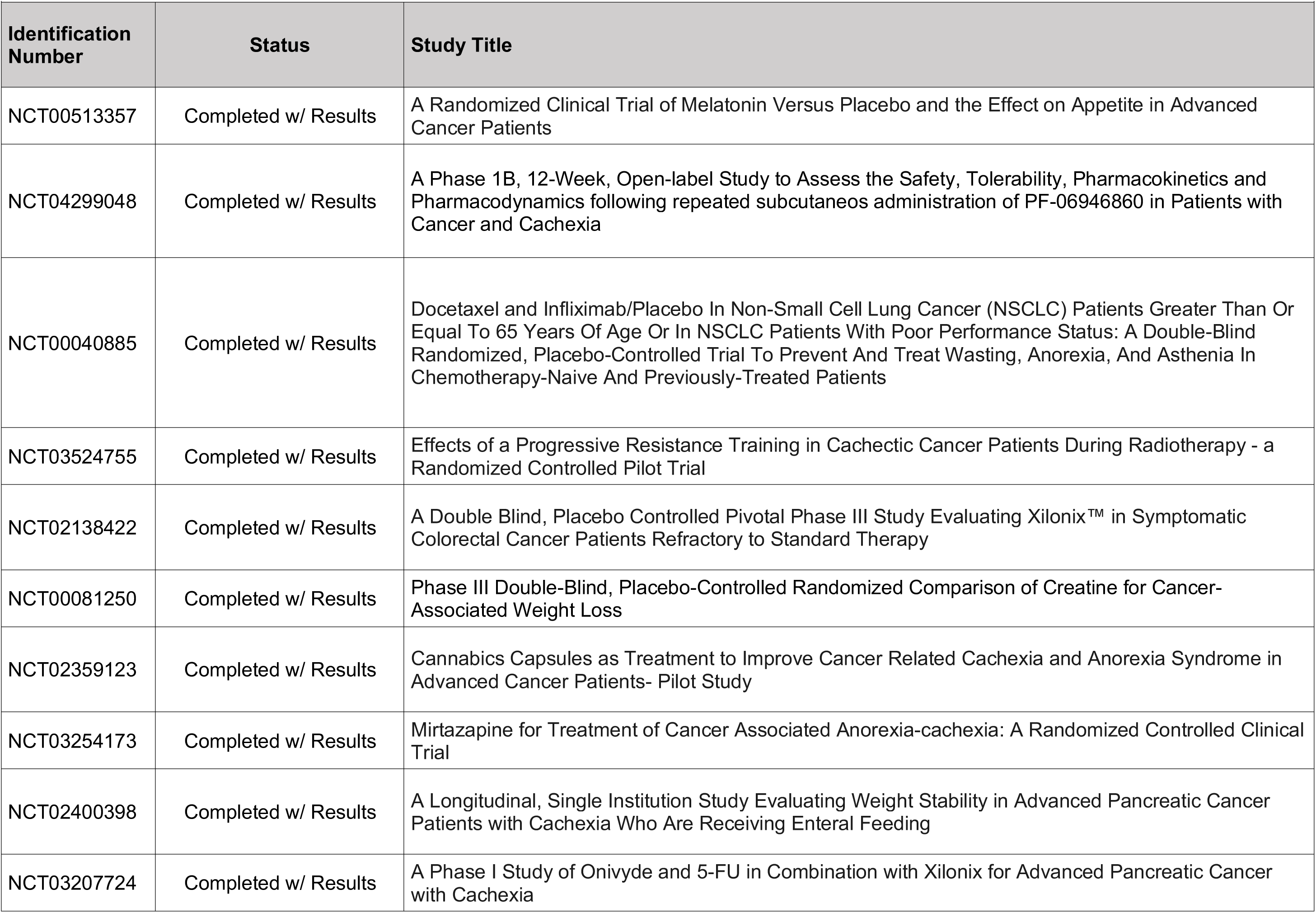
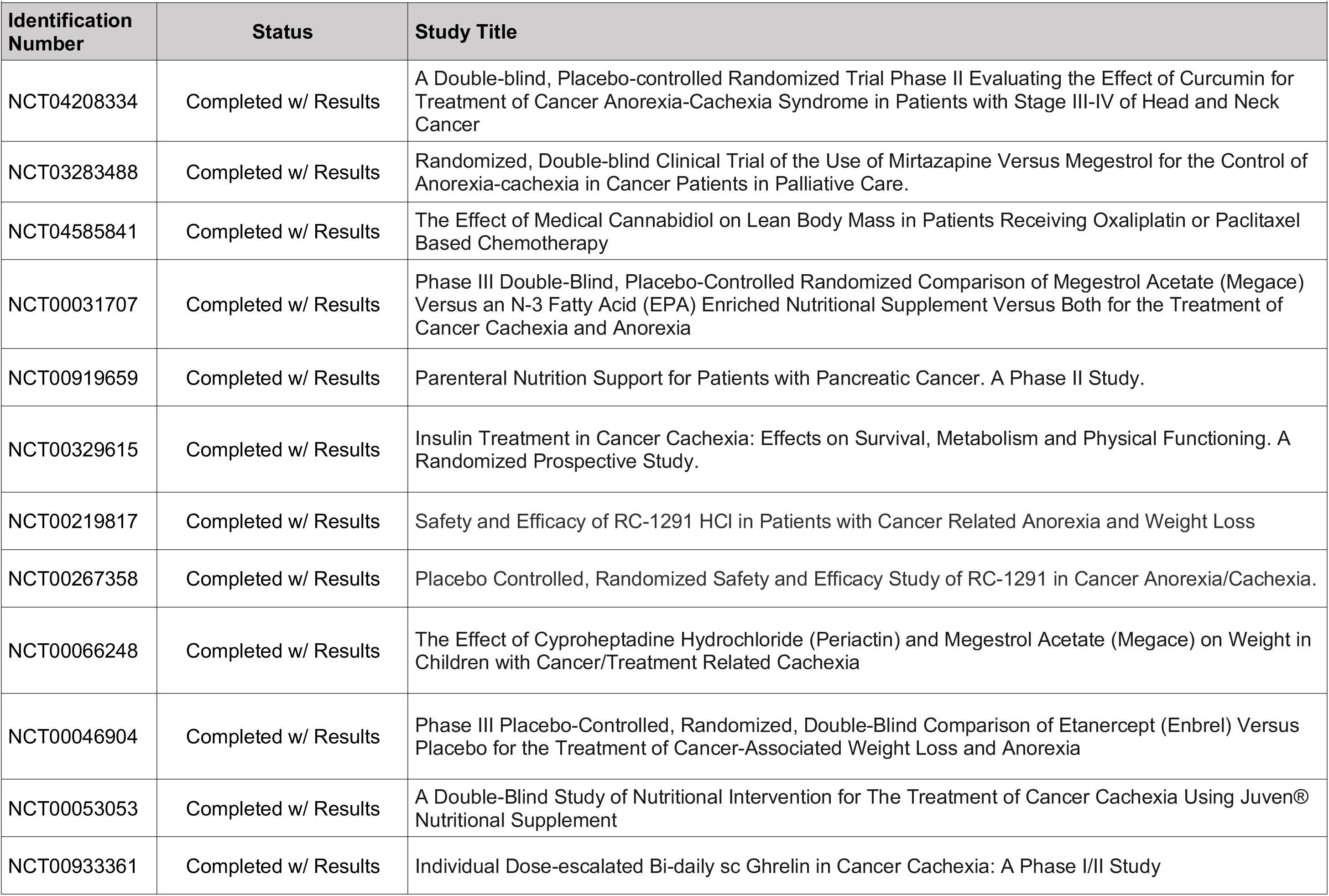
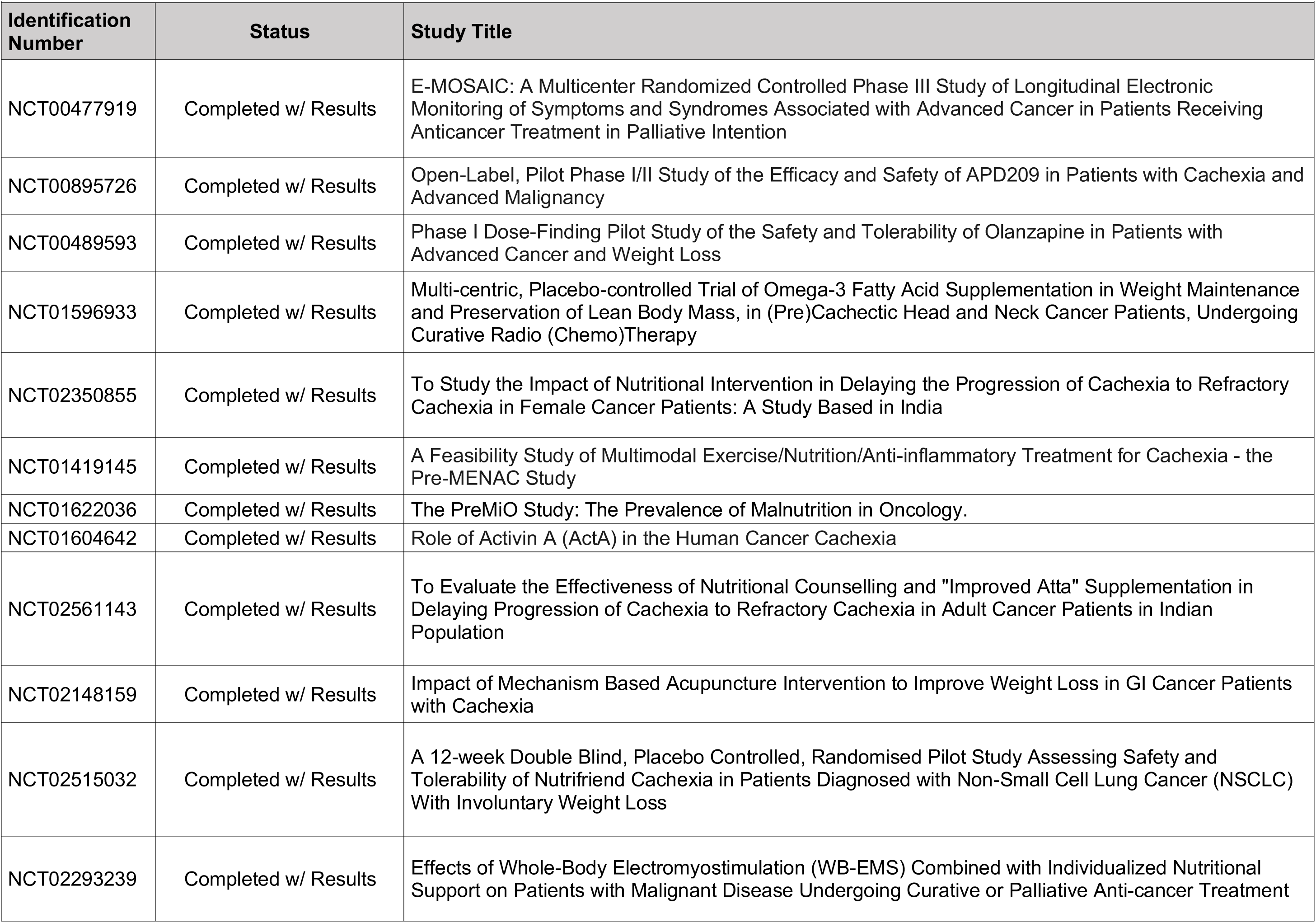
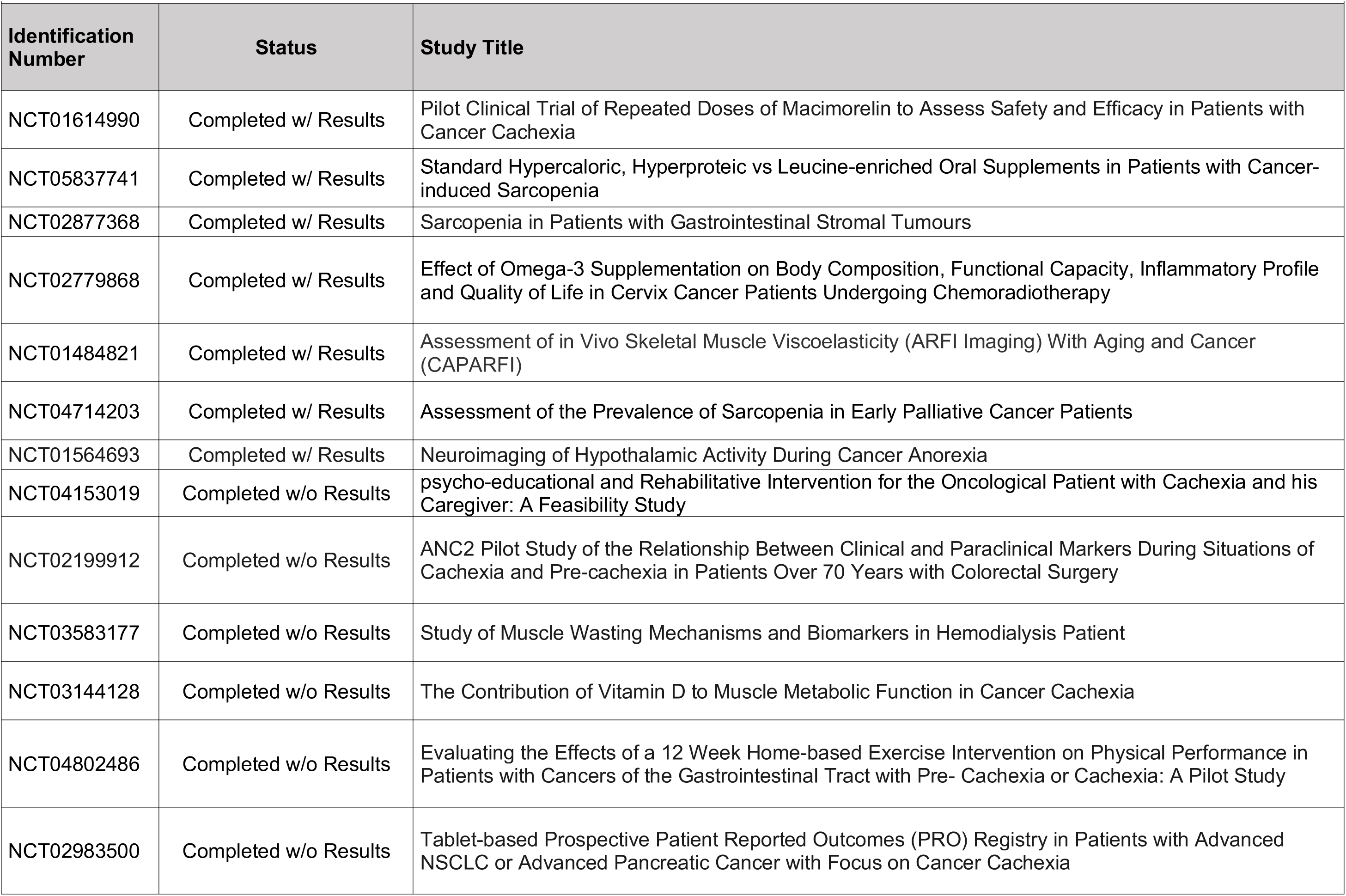
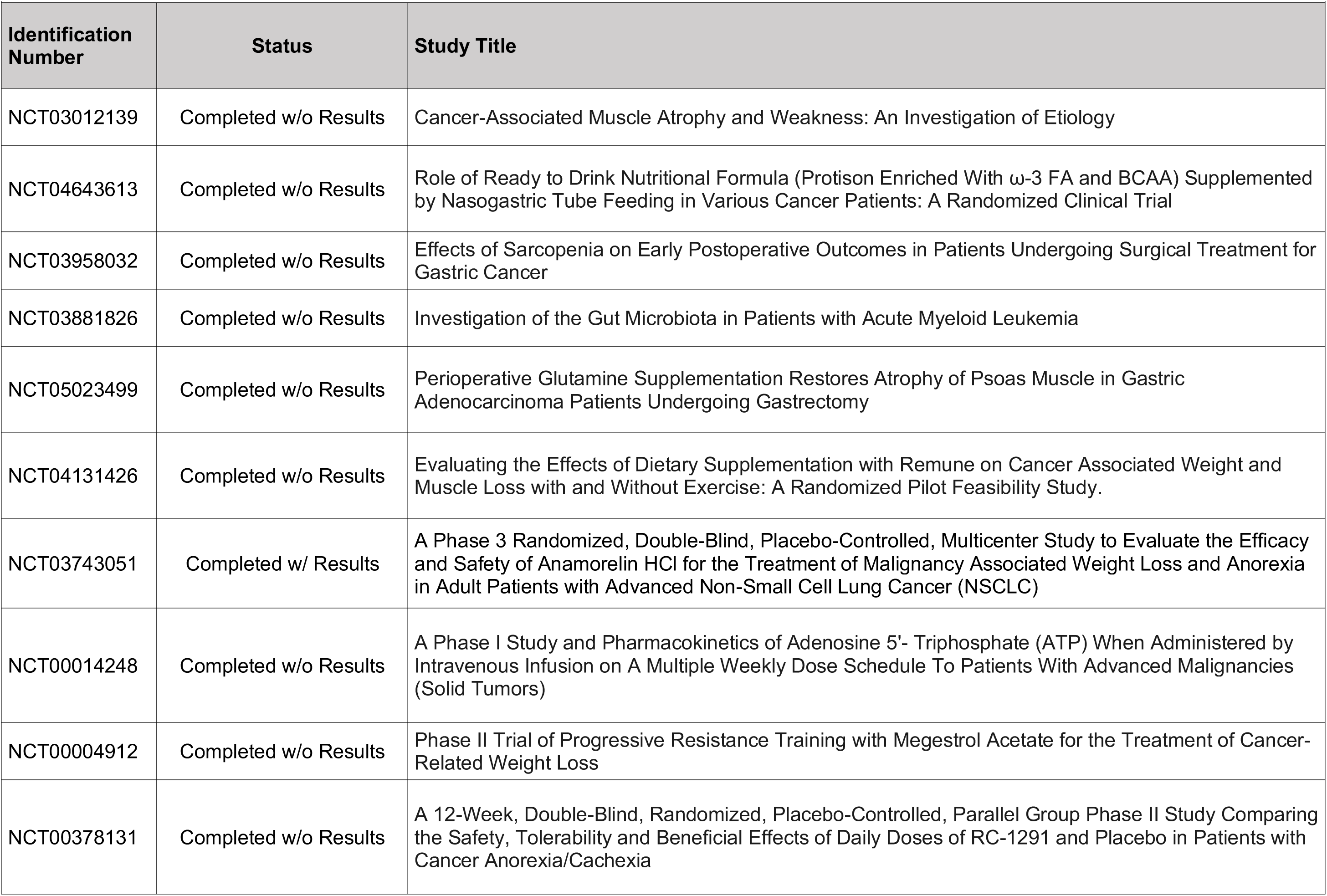
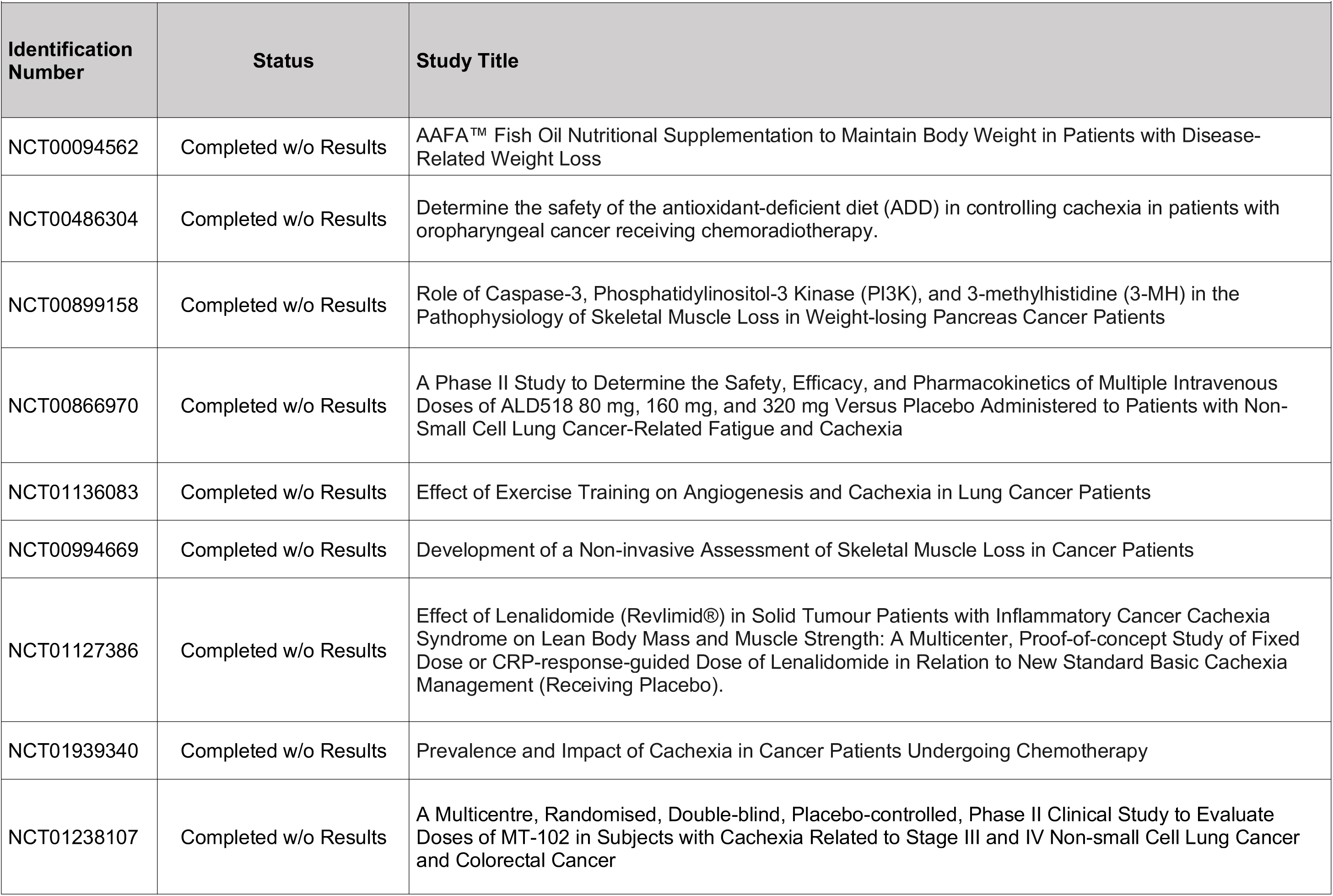
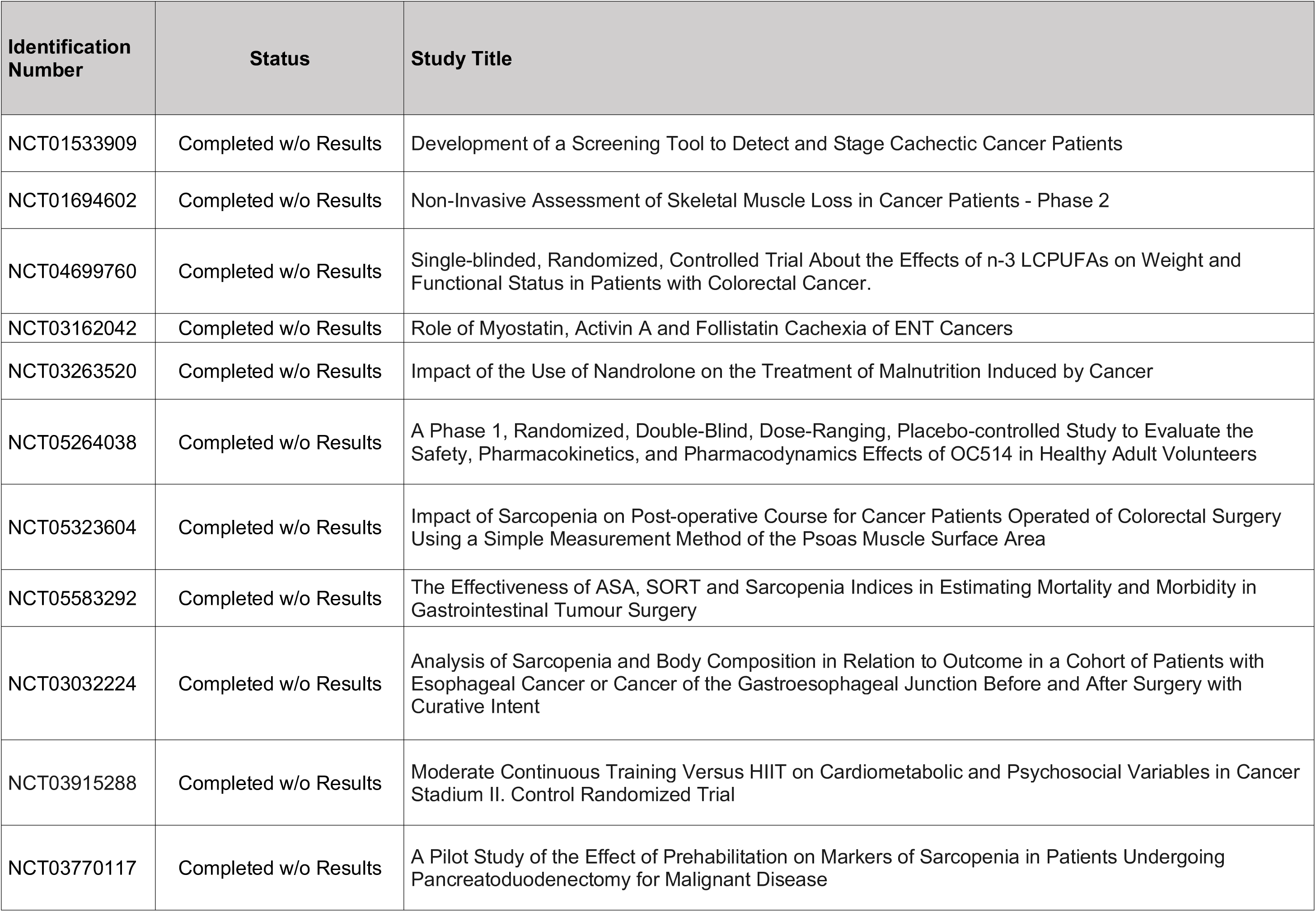
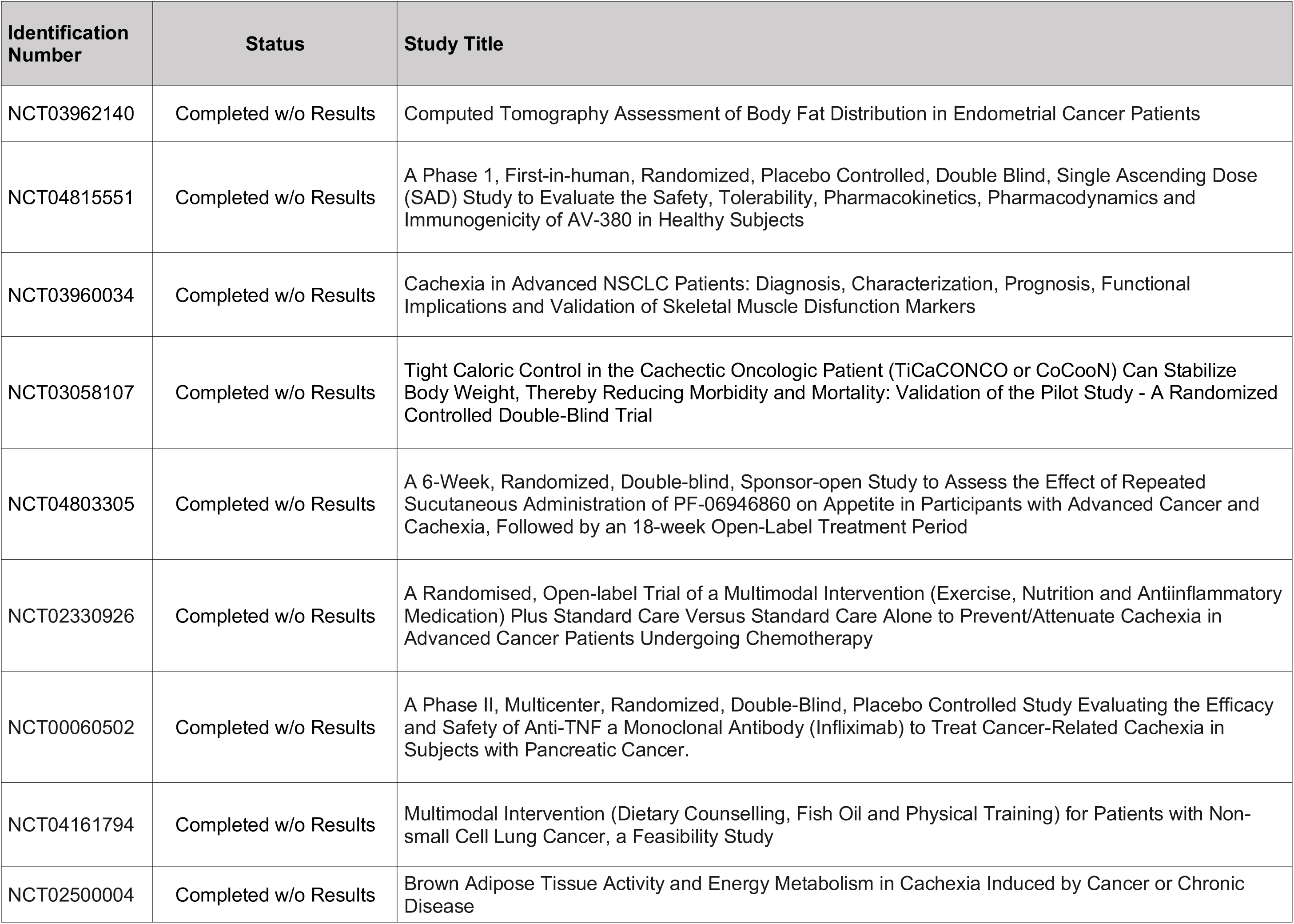
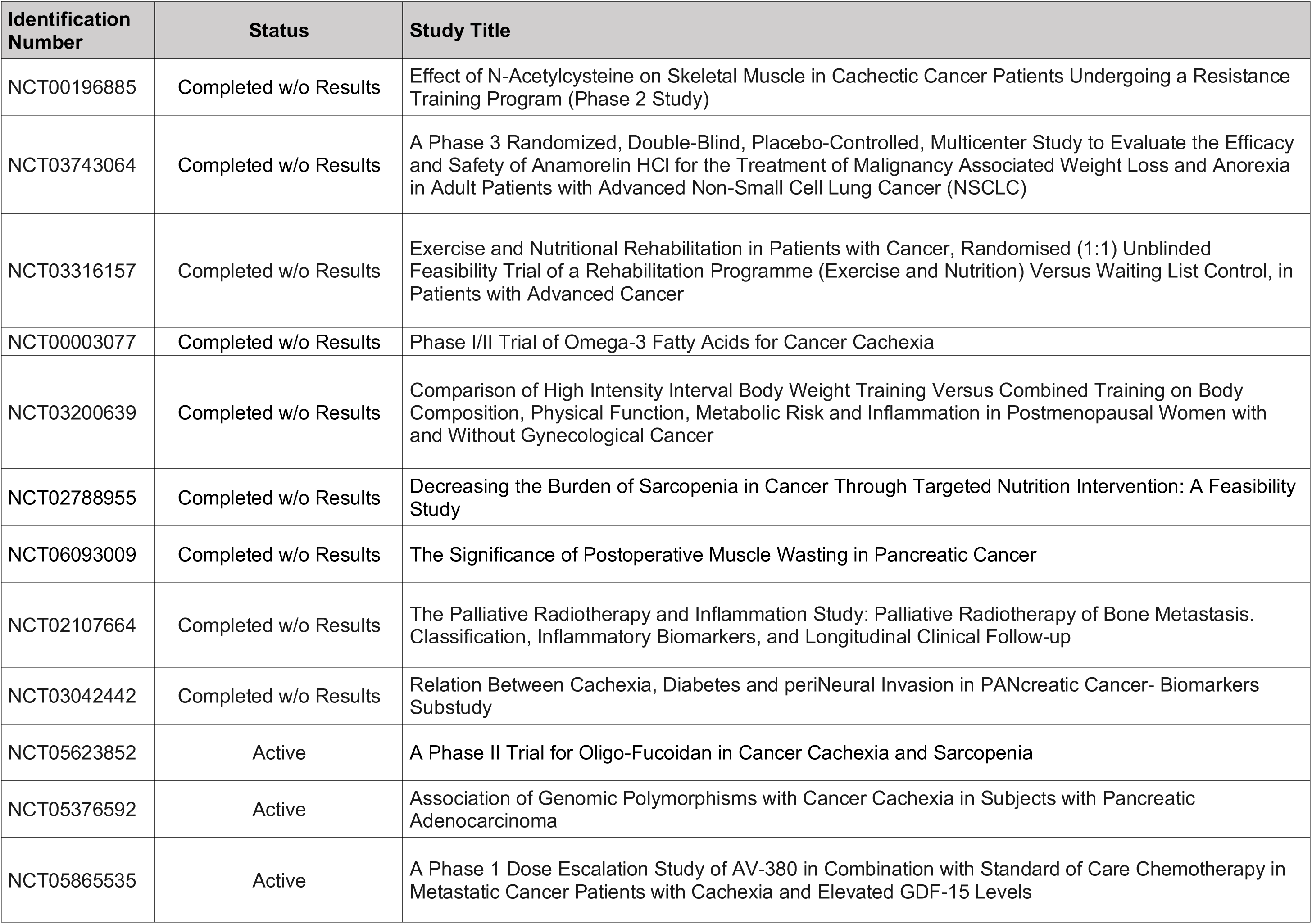
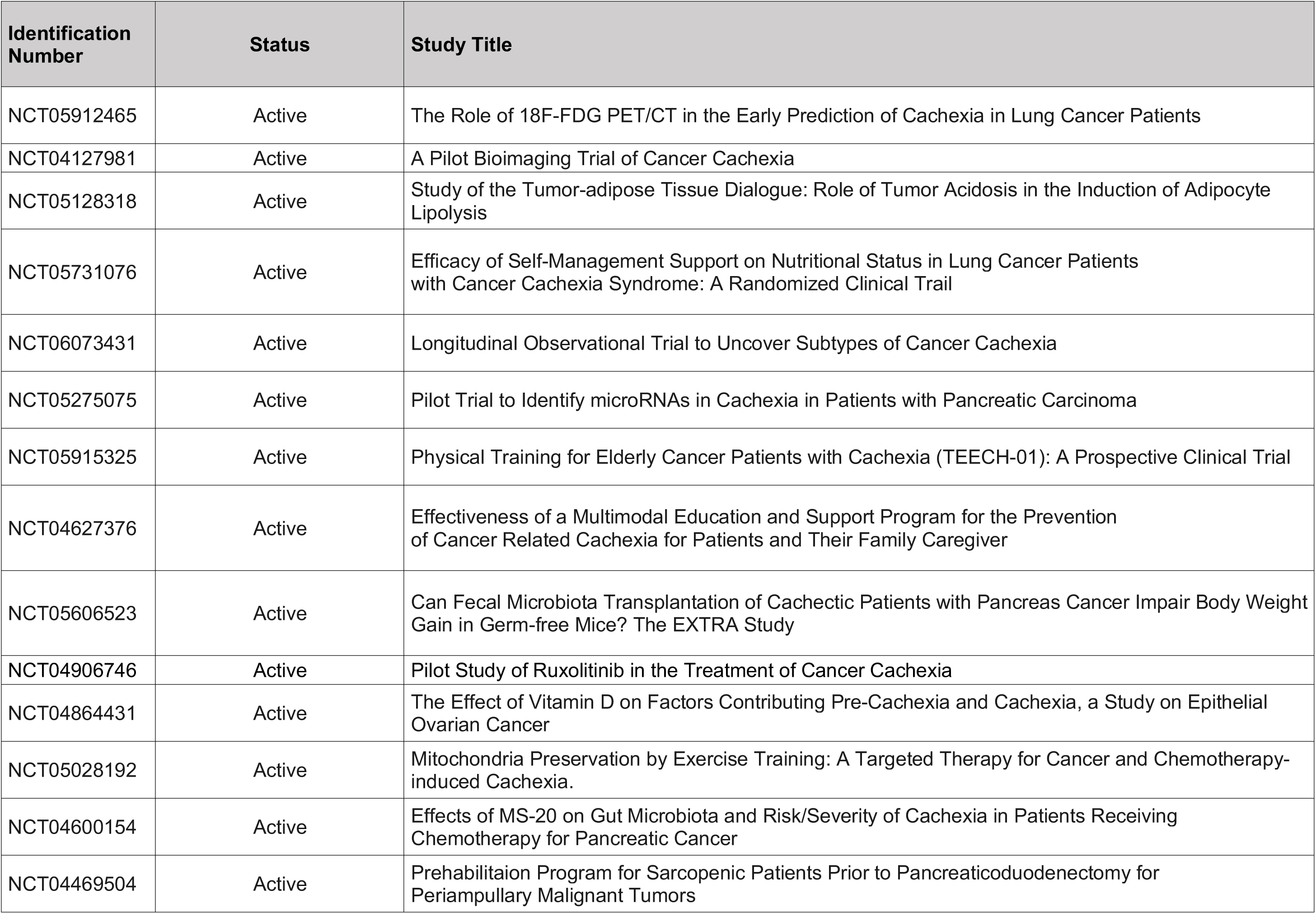
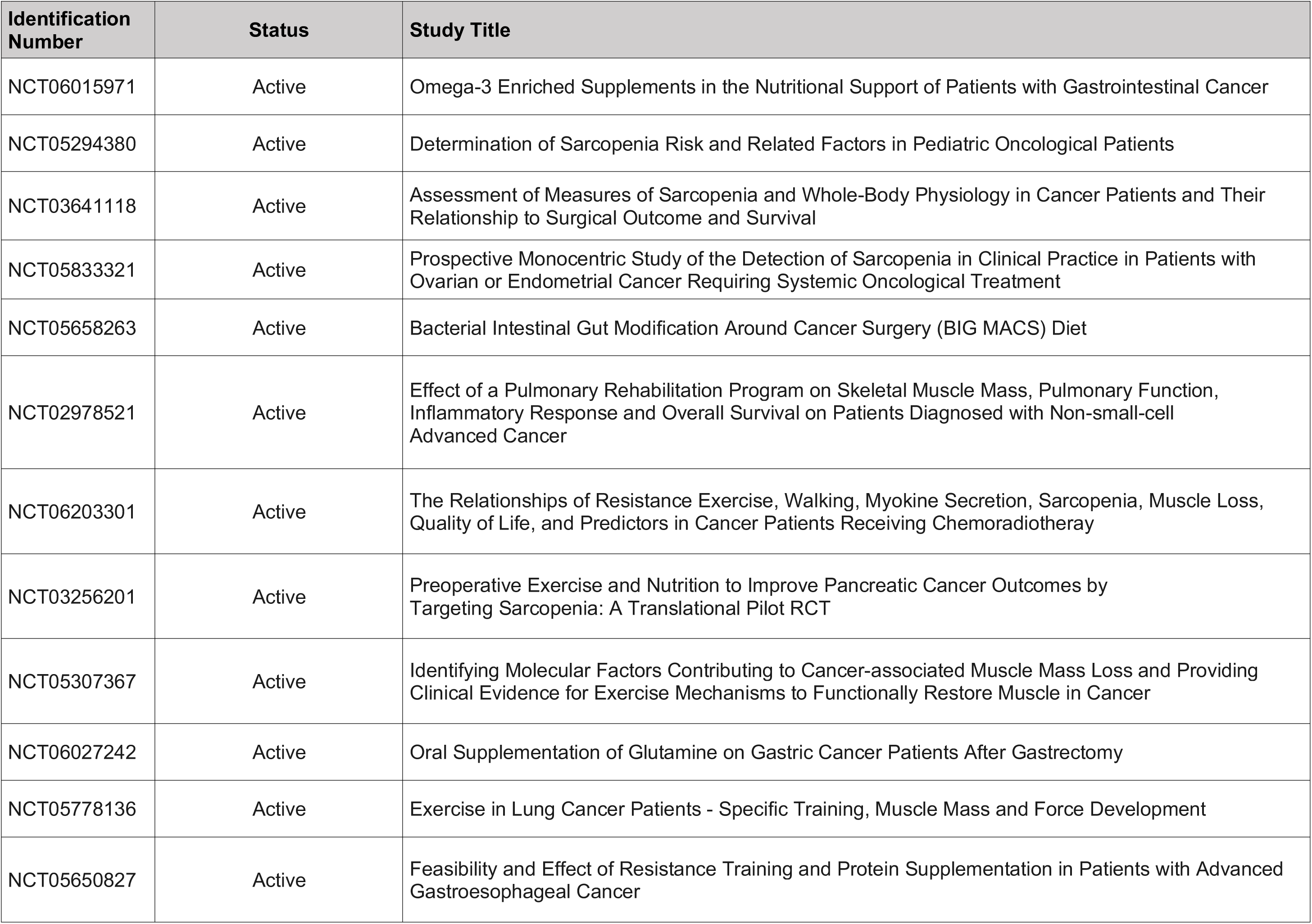
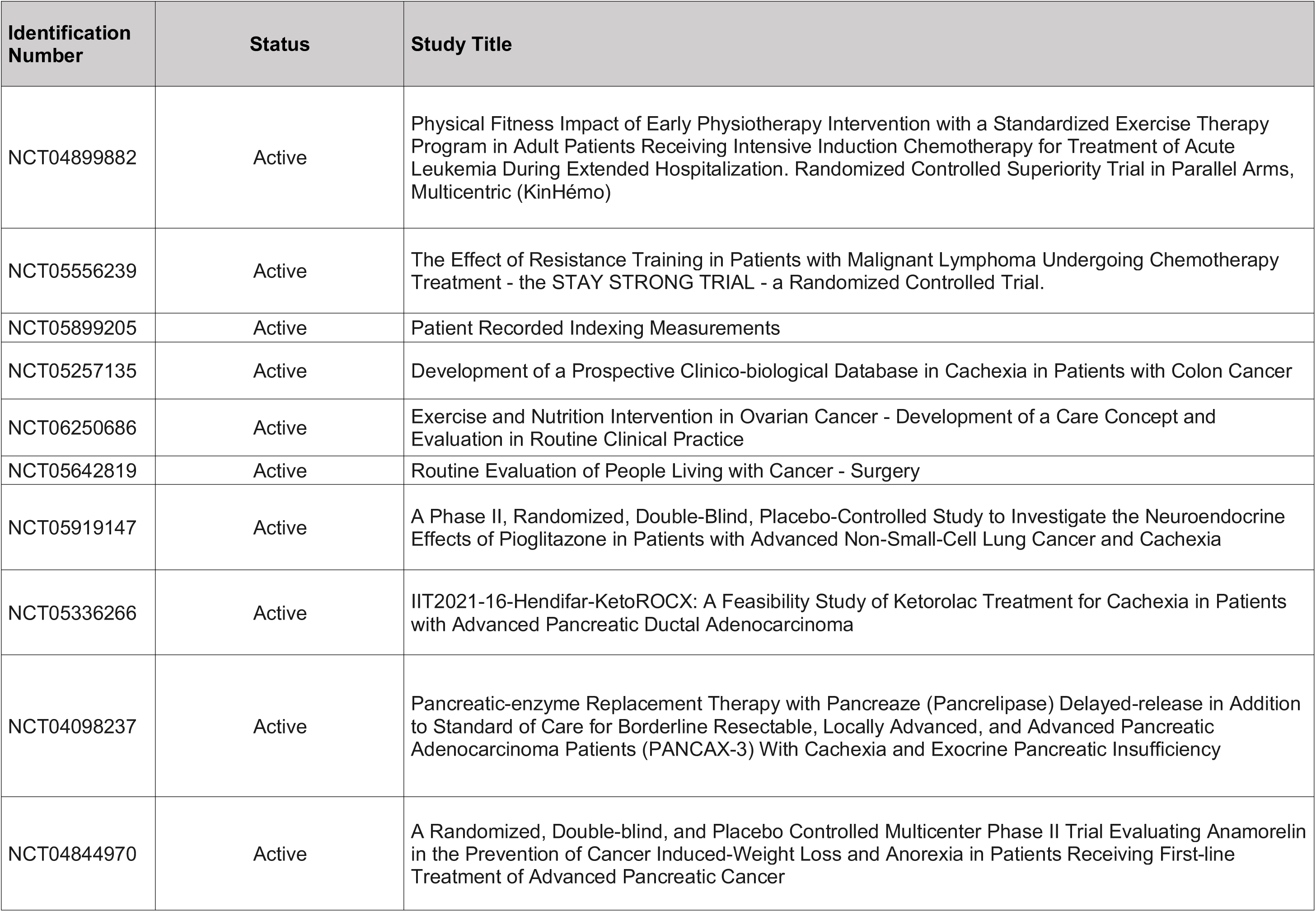
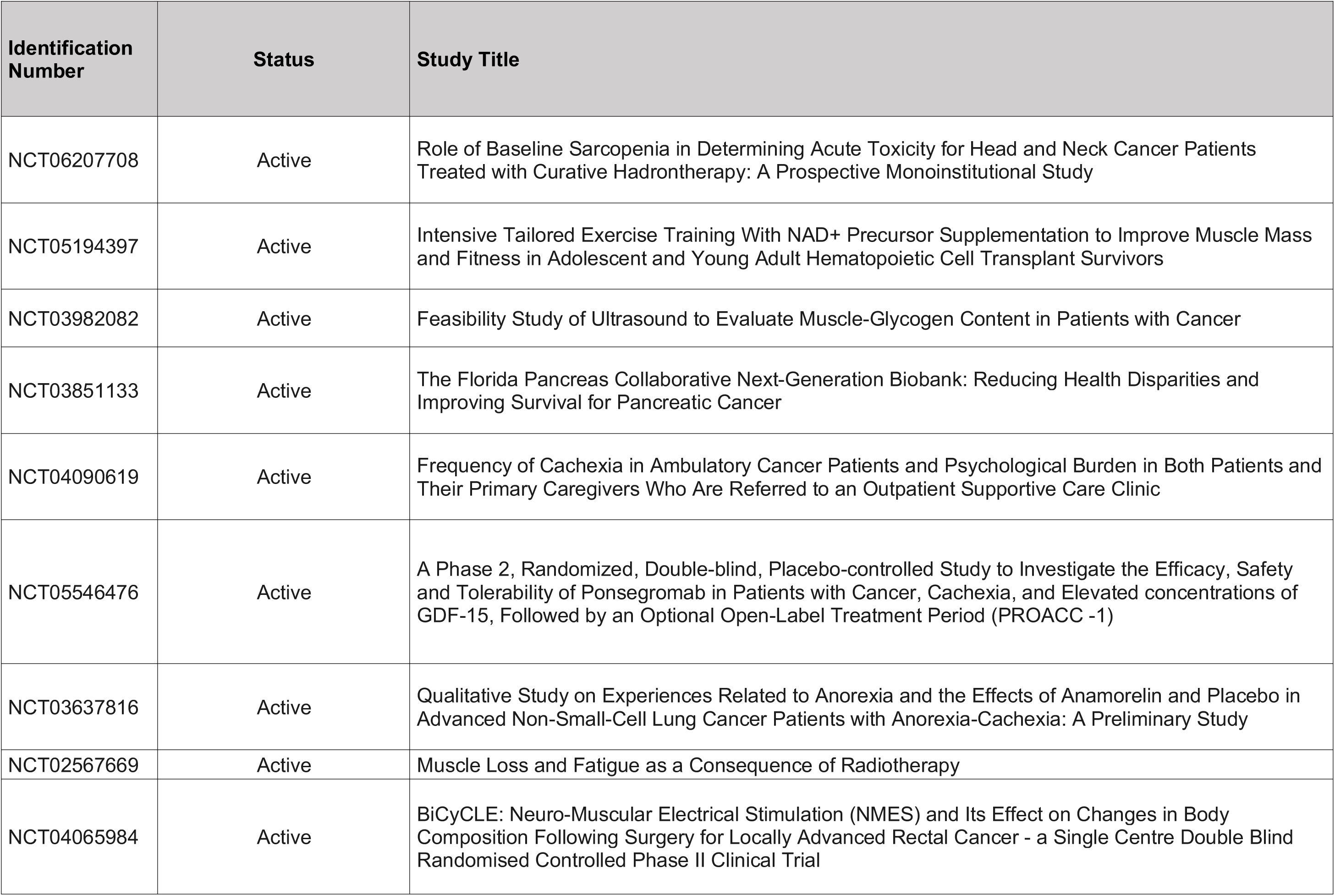
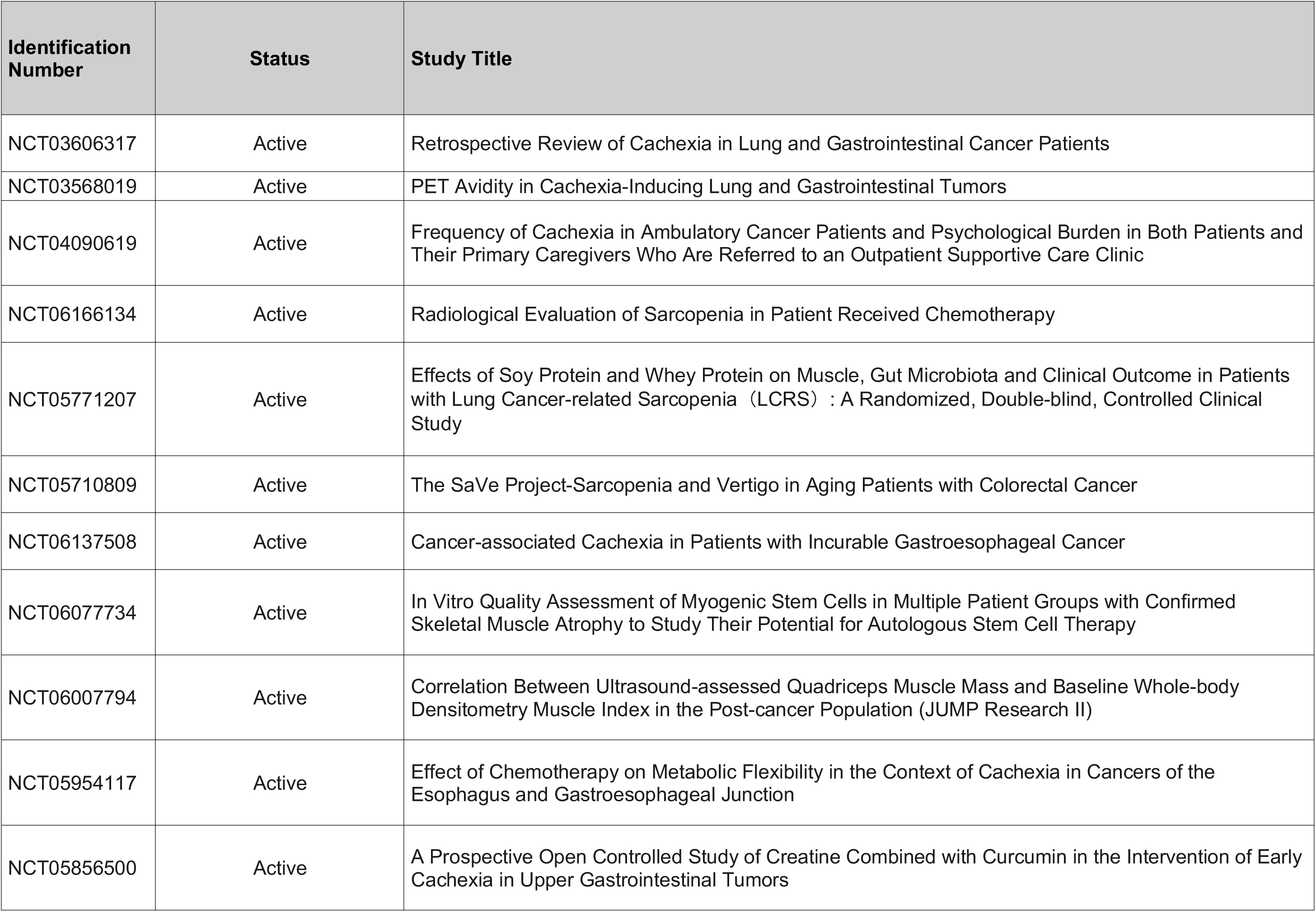
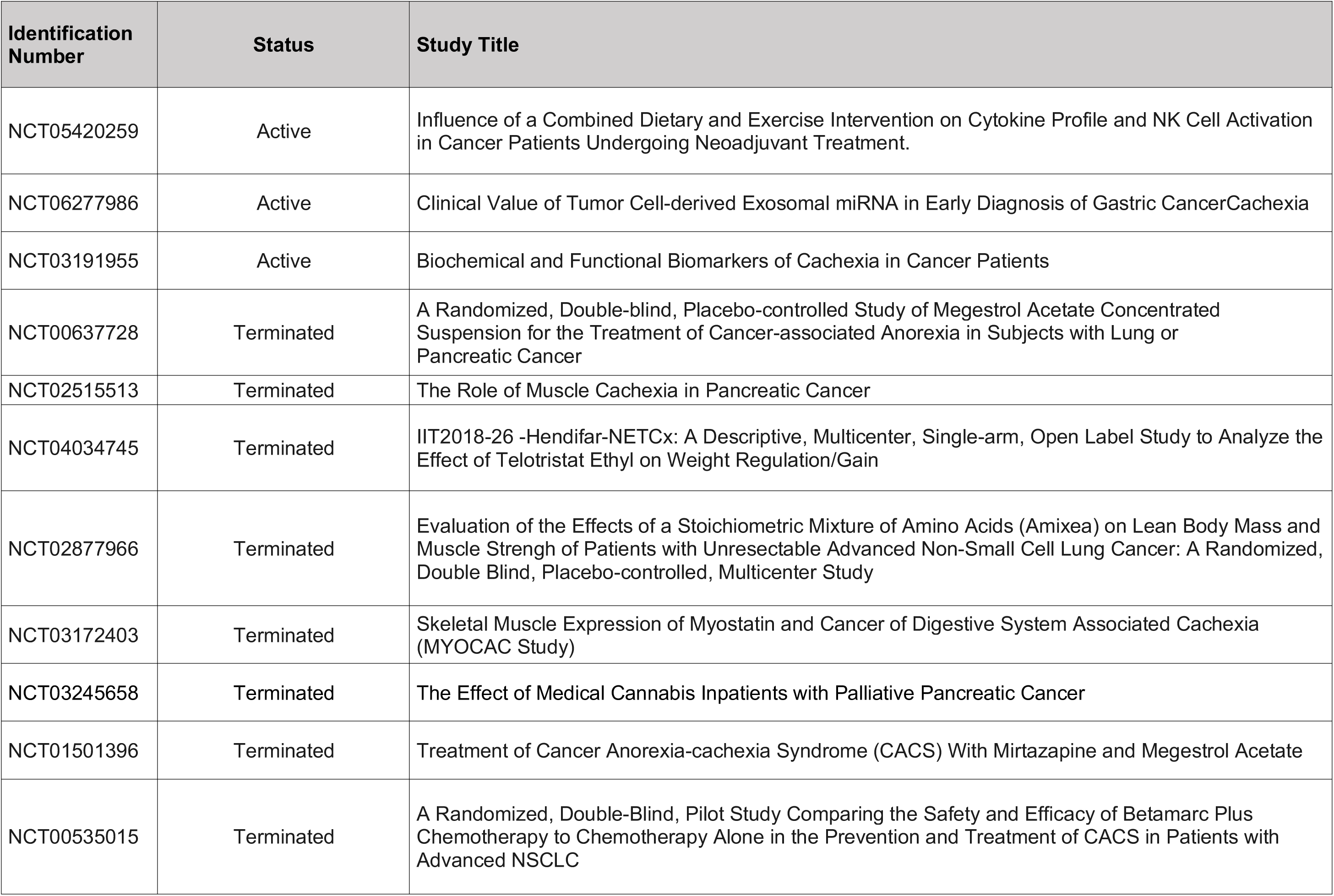
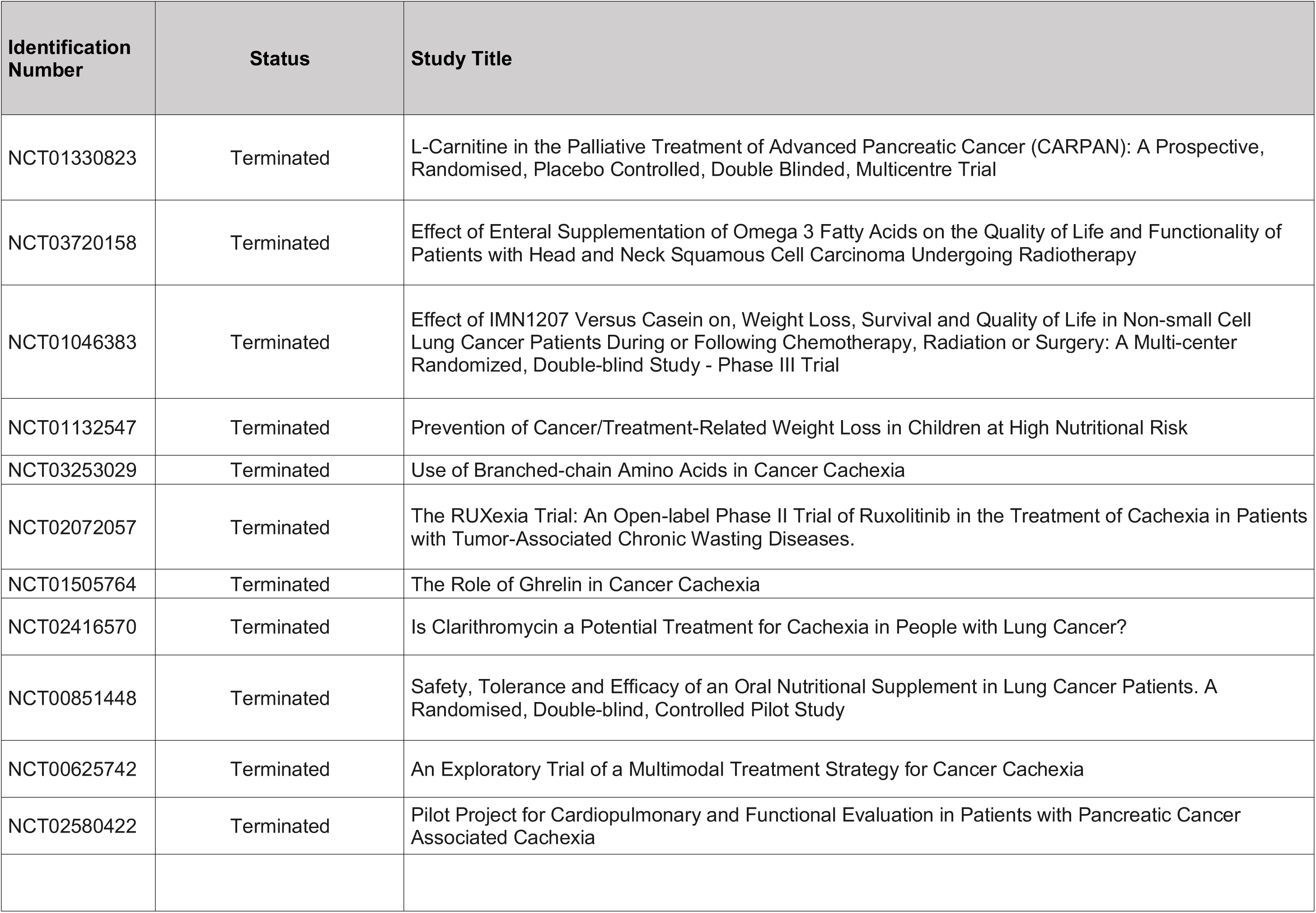
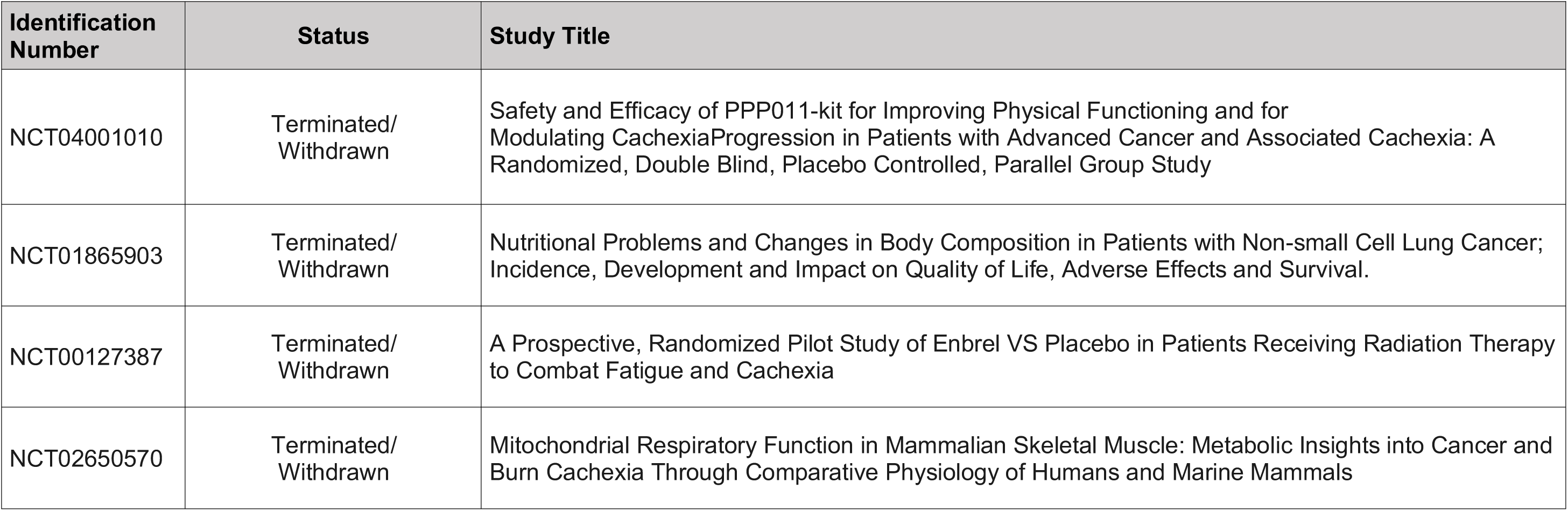
List of clinical trials included in this review. Identification number is the NCT code under which the clinical trial is registered in ClinicalTrials.gov. The status is completed with (w/) results, completed without (w/o) results, still active, or withdrawn (terminated) before completion.

**Supplemental File 2.**
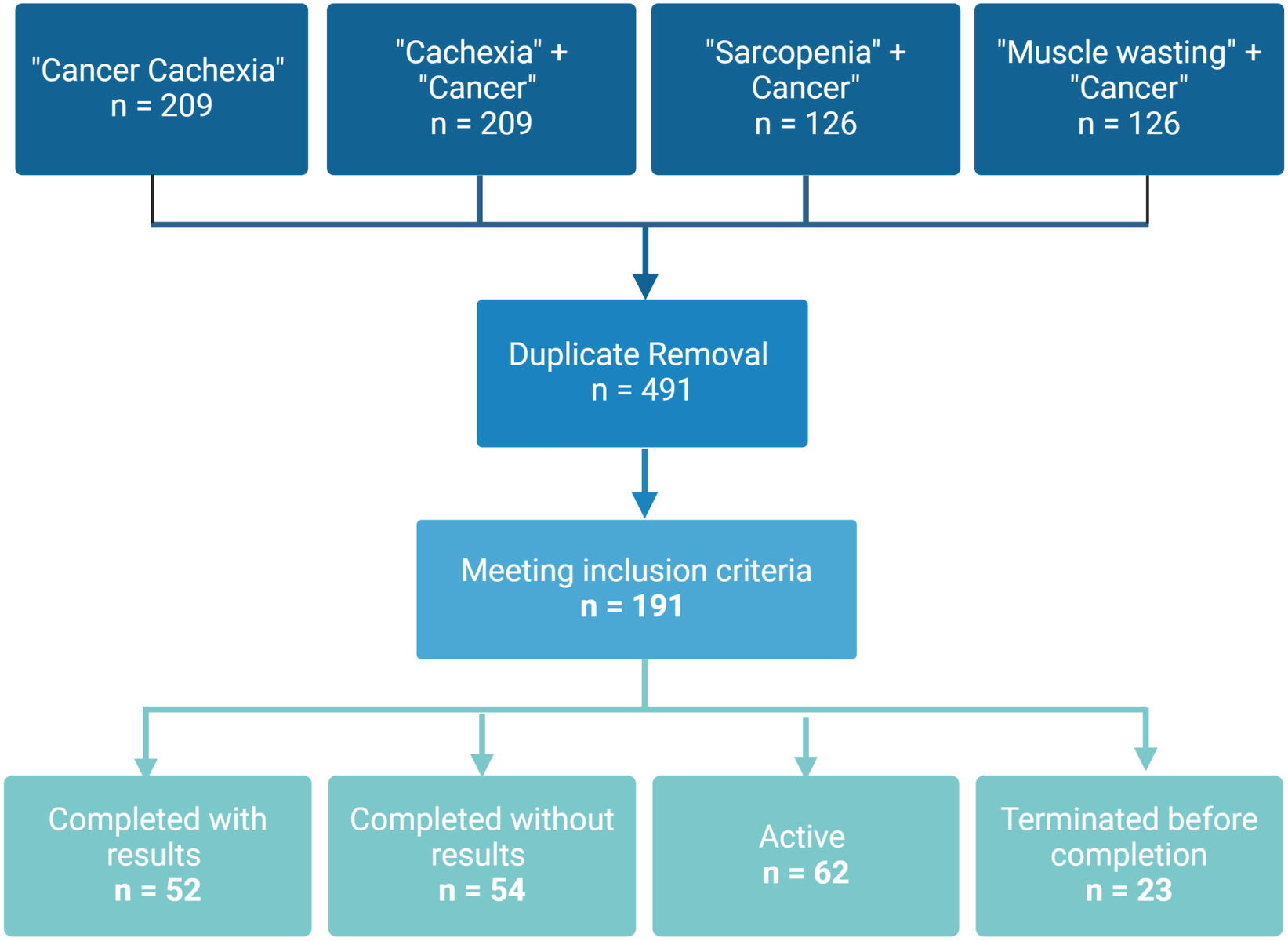
Summary of search results by terms and status of the clinical trials.

**Supplemental File 3.** This table list the reasons for withdrawn or anticipated termination before completion of clinical trials studying cancer cachexia registered in ClinicalTrials.gov.

| Reason for Termination | Trials |
| --- | --- |
|  | n (%) |
| Recruitment | 10 (43.5) |
| Institutional Support & Staff Relocation | 4 (17.4) |
| Funding | 2 (8.7) |
| Change of Sponsor | 1 (4.3) |
| Data variation | 1 (4.3) |
| Time | 1 (4.3) |
| Covid-19 | 1 (4.3) |
| Not Specified | 3 (13.0) |

**Supplemental File 4.** Phases and status of clinical trials exploring cancer cachexia effects on ClinicalTrials.gov in Phase1-4 or No Phase.

| Status/Phase | Phase 1 | Phase 2 | Phase 3 | Phase 4 | No Phase | Other* |
| --- | --- | --- | --- | --- | --- | --- |
| **Completed With Results** | 5 | 12 | 12 | 1 | 19 | 3 |
| **Completed Without Results** | 6 | 8 | 3 | 1 | 34 | 2 |
| **Active** | 4 | 5 | 0 | 0 | 52 | 1 |
| **Withdrawn** | 0 | 6 | 5 | 0 | 11 | 1 |
| **Total** | 15 | 31 | 20 | 2 | 116 | 7 |

Numbers represent the amount of clinical trials registered on ClinicalTrials.gov in Phase1-4 or No Phase and their status: completed with results, completed without results, active, or withdrawn. \*One trial completed with results and two completed without results were in phase 1 and 2. Two trials completed with results, one active, and one withdrawn were in phase 2 and 3.

**Supplemental File 5.** Clinical trials registered on ClinicalTrials.gov that used Fearon’s definition or any other criteria to diagnose cancer cachexia within each status.

| Definition used | Completed with Results (n=52) |  | Completed Without Results (n=54) |  | Active (n=62) |  | Withdrawn (n=23) |  | Total (n=191) |  |
| --- | --- | --- | --- | --- | --- | --- | --- | --- | --- | --- |
|  | n | % | n | % | n | % | n | % | n | % |
| Fearon | 29 | (55.8) | 9 | (16.6) | 24 | (38.7) | 2 | (8.7) | 64 | (33.5) |
| Other | 14 | (26.9) | 19 | (35.2) | 8 | (12.9) | 13 | (56.5) | 54 | (28.3) |
| Not Specified | 9 | (17.3) | 26 | (48.2) | 30 | (48.4) | 8 | (34.8) | 73 | (38.2) |

n represents the amount of clinical trials registered on ClinicalTrials.gov that used Fearon's definition or any other criteria to diagnose cancer cachexia. % refers to the percentage of clinical trials relative the total of clinical trials within each status (completed with results, completed without results, active, or withdrawn). The % in the "Total" column shows the percentage relative to the total of trials included in the study.

**Supplemental File 6.** Clinical trials stratified by clinical trial status studying cachexia but classified under other terms different than “cachexia”.

| Other Terms<br>Used for Trial<br>Classification | Completed<br>with Results<br>(n=52) |  | Completed<br>Without<br>Results<br>(n=54) |  | Active<br>(n=62) |  | Withdrawn<br>(n=23) |  | Total<br>(n=191) |  |
| --- | --- | --- | --- | --- | --- | --- | --- | --- | --- | --- |
|  | n | % | n | % | n | % | n | % | n | % |
| Sarcopenia | 5 | (9.6) | 6 | (11.1) | 16 | (25.8) | 0 | (0.0) | 27 | (14.1) |
| Weight loss | 9 | (17.3) | 10 | (18.5) | 1 | (1.6) | 3 | (13.0) | 23 | (12.0) |
| Muscle loss | 4 | (7.7) | 6 | (11.1) | 9 | (14.5) | 1 | (4.4) | 20 | (10.5) |
| Malnutrition | 4 | (7.7) | 3 | (5.5) | 1 | (1.6) | 1 | (4.4) | 9 | (4.7) |

n represents the number trials classified as sarcopenia, weight loss, muscle loss, or malnutrition studies. Trials are stratified by clinical trial status (completed with results, completed without results, active, or withdrawn).

**Supplemental File 7.** **A)** Study population of clinical trials registered on ClinicalTrials.gov investigating cancer cachexia. **B)** Population dropout percentage within each clinical trial.

| Study Population |  | Completed Trials with Results (n=52) | Completed Trials Without Results (n=54) | Active Trials (n=62) | Withdrawn Trials (n=23) | Total (n=191) |
| --- | --- | --- | --- | --- | --- | --- |
|  |  | n (%) | n (%) | n (%) | n (%) | n (%) |
| Enrollment | 1-50 | 22 (42.3) | 19 (35.2) | 18 (29.0) | 13 (56.5) | 72 (37.7) |
|  | 51-100 | 12 (23.1) | 17 (31.5) | 18 (29.0) | 2 (8.7) | 49 (25.7) |
|  | 101-500 | 15 (28.8) | 13 (24.1) | 22 (35.5) | 1 (4.4) | 51 (26.7) |
|  | 501-1000 | 2 (3.8) | 2 (3.7) | 2 (3.2) | 0 (0.0) | 6 (3.1) |
|  | >1000 | 1 (1.9) | 1 (1.9) | 2 (3.2) | 0 (0.0) | 4 (2.1) |
|  | Not specified | 0 (0.0) | 2 (3.7) | 0 (0.0) | 7 (30.4) | 9 (4.7) |

n represents the number of trials by range of participants enrolled to the clinical trials by clinical trial status (completed with results, completed without results, active, or withdrawn). % refers to the percentage of clinical trials relative the total of clinical trials within each status. The % in the “Total” column shows the percentage relative to the total of trials included in the study.
Dropout percentage (%) in the CTx completed with results that reported “Started” and “Not completed” populations. The dropout percentage is relative to each trial’s population (n=28). The “Started” population is labeled on each bar.

**Supplemental File 8.** Cancer type and cancer stage of clinical trials registered on ClinicalTrials.gov investigating cancer cachexia.

| Cancer Type and Stage |  | Completed Trials with Results (n=52) | Completed Trials Without Results (n=54) | Active Trials (n=62) | Withdrawn Trials (n=23) | Total (n=191) |
| --- | --- | --- | --- | --- | --- | --- |
|  |  | n (%) | n (%) | n (%) | n (%) | n (%) |
| Type of Cancer Studied | **Single cancer type** |  |  |  |  |  |
|  | Lung | 8 (15.4) | 6 (11.1) | 10 (16.1) | 5 (21.7) | 29 (15.2) |
|  | Pancreatic | 3 (5.8) | 4 (7.4) | 11 (17.7) | 4 (17.4) | 22 (11.5) |
|  | Gastric/GI* | 3 (5.8) | 5 (9.3) | 8 (12.9) | 1 (4.3) | 17 (8.9) |
|  | Colorrectal | 1 (1.9) | 4 (7.4) | 5 (8.1) | 0 (0.0) | 10 (5.2) |
|  | Head & Neck | 3 (5.8) | 1 (1.9) | 1 (1.6) | 2 (8.7) | 7 (3.7) |
|  | Others | 2 (3.8) | 4 (7.4) | 5 (8.1) | 0 (0.0) | 11 (5.8) |
|  | **Combination of 2** |  |  |  |  |  |
|  | Lung + other | 5 (9.6) | 5 (9.3) | 2 (3.2) | 2 (8.7) | 14 (7.3) |
|  | GI + other | 0 (0.0) | 2 (3.7) | 1 (1.6) | 0 (0.0) | 3 (1.6) |
|  | **Combination of 3** |  |  |  |  |  |
|  | Lung + Pancreatic + Colorectal | 1 (1.9) | 0 (0.0) | 2 (3.2) | 1 (4.3) | 4 (2.1) |
|  | Others | 1 (1.9) | 2 (3.7) | 2 (3.2) | 1 (4.3) | 6 (3.1) |
|  | **Combination of 4 or more** | 8 (15.4) | 6 (11.1) | 6 (9.7) | 2 (8.7) | 22 (11.5) |
|  | **Not specified** | 17 (32.7) | 15 (27.8) | 9 (14.5) | 5 (21.7) | 46 (24.1) |
| Cancer Stage | **Single stage** | 0 (0.0) | 2 (3.7) | 2 (3.2) | 2 (8.7) | 6 (3.1) |
|  | **Mixed** |  |  |  |  |  |
|  | Stage I-III | 0 (0.0) | 0 (0.0) | 1 (1.6) | 0 (0.0) | 1 (0.5) |
|  | Stage II-III | 2 (3.8) | 0 (0.0) | 2 (3.2) | 0 (0.0) | 4 (2.1) |
|  | Stage I-IV | 0 (0.0) | 1 (1.9) | 1 (1.6) | 1 (4.3) | 3 (1.6) |
|  | Stage II-IV | 1 (1.9) | 2 (3.7) | 3 (4.8) | 1 (4.3) | 7 (3.7) |
|  | Stage III-IV | 9 (17.3) | 5 (9.3) | 4 (6.5) | 6 (26.1) | 24 (12.6) |
|  | **Not Specified** | 40 (76.9) | 44 (81.5) | 49 (79.0) | 13 (56.5) | 146 (76.4) |

n represents the number of clinical trials that stated 1, 2, 3, or 4 and more cancer types and the cancer stage. % refers to the percentage of clinical trials relative the total of clinical trials within each status. The % in the "Total" column shows the percentage relative to the total of trials included in the study. \*GI = gastrointestinal. Due to the uncertainty in some studies that reported gastric or gastrointestinal cancer without further details, gastrointestinal cancer was considered as a singular type of cancer rather than as a collection of conditions that can affect the entire gastrointestinal tract.

**Supplemental File 9.** Clinical trials approach and intervention utilized: drug testing, dietary supplement, exercise/physical therapy, a combination of 2 or more interventions (multimodal), others, or none or non-stated intervention of clinical trials registered on ClinicalTrials.gov investigating cancer cachexia.

| Approach |  | Completed<br>with Results<br>(n=52) | Completed<br>Without<br>Results<br>(n=54) | Active<br>(n=62) | Withdrawn<br>(n=23) | Total<br>(n=191) |
| --- | --- | --- | --- | --- | --- | --- |
|  |  | n % | n % | n % | n % | n % |
| Study<br>Type | Observational | 7 (13.5) | 20 (37.0) | 20 (32.3) | 4 (17.4) | 51 (26.7) |
|  | Interventional | 45 (86.5) | 34 (63.0) | 42 (67.7) | 19 (82.6) | 140 (73.3) |
| Intervention | Drug | 28 (53.8) | 14 (25.9) | 11 (17.7) | 11 (47.8) | 64 (33.5) |
|  | Dietary Supplement | 8 (15.4) | 7 (13.0) | 7 (11.3) | 6 (26.0) | 28 (14.7) |
|  | Exercise/Physical Therapy | 1 (1.9) | 6 (11.1) | 8 (12.9) | 0 (0.0) | 15 (7.9) |
|  | Multimodal | 6 (11.5) | 3 (5.6) | 8 (12.9) | 1 (4.4) | 18 (9.4) |
|  | Dietary Supplement and Exercise | 5 (9.6) | 0 (0.0) | 2 (3.2) | 0 (0.0) | 7 (3.7) |
|  | Dietary Supplement and Drug | 1 (1.9) | 0 (0.0) | 0 (0.0) | 0 (0.0) | 1 (0.5) |
|  | Dietary Supplement and Nutrition Counseling | 0 (0.0) | 1 (1.9) | 0 (0.0) | 0 (0.0) | 1 (0.5) |
|  | Dietary Supplement, Exercise, and Drug | 0 (0.0) | 0 (0.0) | 0 (0.0) | 1 (4.4) | 1 (0.5) |
|  | Drug and Exercise | 0 (0.0) | 2 (3.7) | 0 (0.0) | 0 (0.0) | 2 (1.1) |
|  | Nutrition Counseling and Exercise | 0 (0.0) | 0 (0.0) | 4 (6.5) | 0 (0.0) | 4 (2.1) |
|  | Nutrition Counseling, Exercise, and Psychological | 0 (0.0) | 0 (0.0) | 1 (1.6) | 0 (0.0) | 1 (0.5) |
|  | Education and Psychological | 0 (0.0) | 0 (0.0) | 1 (1.6) | 0 (0.0) | 1 (0.5) |
|  | Other | 2 (3.8) | 4 (7.4) | 10 (16.1) | 1 (4.4) | 17 (8.9) |
|  | None | 7 (13.5) | 20 (37.0) | 18 (29.0) | 4 (17.4) | 49 (25.7) |

n represents the amount of clinical trials registered on ClinicalTrials.gov that utilized and observational or interventional approach for their study. This table also shows the intervention utilized (drug testing, dietary supplement, exercise/physical therapy, a combination of 2 or more interventions (multimodal), others, or none or non-stated intervention. % refers to the percentage of clinical trials relative the total of clinical trials within each status (completed with results, completed without results, active, or withdrawn). The % in the “Total” column shows the percentage relative to the total of trials included in the study.

**Supplemental File 10.** Pharmaceutical drugs and dietary supplements evaluated in CTx between 1995-2024.

| CTx Identification Number | Treatment | Class |
| --- | --- | --- |
| NCT00866970 | ALD518 | IL-6 inhibitor / Monoclonal antibody |
| NCT02138422, NCT03207724 | Xilonix | Anti-IL-1α monoclonal antibody |
| NCT01395914, NCT01387282, NCT01387269, NCT03743064, NCT03637816, NCT01505764, NCT04844970, NCT00219817, NCT00267358, NCT00378131 | Anamorelin HCl / RC-1291 | Ghrelin receptor agonist / Appetite stimulant |
| NCT00066248, NCT01132547 | Cyproheptadine hydrochloride | Antihistamine / Appetite stimulant |
| NCT00933361 | Ghrelin | Peptide hormone (appetite stimulant) |
| NCT01614990 | Macimorelin | Ghrelin receptor agonist |
| NCT03283488, NCT00004912, NCT00637728, NCT01501396, NCT00066248 | Megestrol | Progestin / Appetite stimulant |
| NCT00625742 | Atenolol | β-blocker (antihypertensive) |
| NCT01238107 | MT-102 | β-blocker + anabolic agent |
| NCT00895726 | APD209 | β-Adrenoceptor agonist |
| NCT00535015 | Betamarc | β-adrenergic agent |
| NCT04815551, NCT05865535 | AV-380 | Anti-GDF-15 |
| NCT05546476, NCT04299048, NCT04803305 | Ponsegromab / PF-06946860 | Anti-GDF15 |
| NCT01433263 | BYM338 / Bimagrumab | Anti-ActRII |
| NCT02359123 | Cannabics | Cannabis-based therapy |
| NCT04585841 | Cannabidiol (CBD) | Phytocannabinoid |
| NCT03245658 | THC and CBD Mixture | Phytocannabinoids |
| NCT04001010 | PPP011 | Cannabinoid-based |
| NCT06166134, NCT06137508, NCT05954117, NCT00040885, NCT00060502, NCT01127386 | Antineoplastic agents | Chemotherapy / Immunomodulators |
| NCT04600154 | MS-20 | Targeted cancer therapy |
| NCT02416570 | Clarithromycin | Antibiotic |
| NCT00127387, NCT00046904 | Enbrel / Etanercept | TNF inhibitor |
| NCT00040885, NCT00060502 | Infliximab | TNF inhibitor |
| NCT00329615 | Insulatard, flexpen | Long-acting insulin |
| NCT05919147 | Pioglitazone | Thiazolidinedione (insulin sensitizer) |
| NCT03254173, NCT03283488 | Mirtazapine / Remeron | Antidepressant / Appetite stimulant |
| NCT00489593 | Olanzapine | Antipsychotic |
| NCT03263520 | Nandrolone Decanoate; Dexamethasone | Anabolic steroid + Corticosteroid |
| NCT00878995 | Testosterone Enanthate | Anabolic androgenic steroid |
| NCT01355497, NCT01355484, NCT00467844 | GTx-024 / Enobosarm | Selective androgen receptor modulator (SARM) |
| NCT04906746, NCT02072057 | Ruxolitinib | JAK1/2 inhibitor |
| NCT04034745 | Telotristat ethyl | Tryptophan hydroxylase inhibitor |
| NCT00527319 | VT-122 | COX-2 inhibitor + PPARγ agonist |
| NCT05336266 | Ketorolac<br>Tromethamine | Non-steroidal anti-inflammatory |
| NCT00196885 | N-acetylcysteine | Antioxidant / Glutathione precursor |
| NCT00513357, NCT00625742 | Melatonin | Hormone / Sleep aid / Antioxidant |
| NCT04208334, NCT05856500 | Curcumin | Anti-inflammatory / Antioxidant |
| NCT00815685, NCT00031707 | Eicosapentaenoic Acid | ω-3 fatty acid / Anti-inflammatory |
| NCT00094562, NCT04699760, NCT04161794 | Fish oil supplement | ω-3 fatty acid / Anti-inflammatory |
| NCT01596933 | BioMega SDA® | ω-3 fatty acid / Anti-inflammatory |
| NCT03720158, NCT06015971, NCT02779868, NCT00003077 | Omega 3 | ω-3 fatty acid / Anti-inflammatory |
| NCT05623852 | Fucoidan | Immune-modulatory / Antioxidant |
| NCT00014248 | Adenosine triphosphate | Cellular energy molecule |
| NCT00081250, NCT05856500 | Creatine monohydrate | Ergogenic aid |
| NCT02561143 | Atta | Nutritional supplement |
| NCT00625742, NCT00053053 | Juven | Nutritional supplement |
| NCT02515032 | Nutrifriend Cachexia | Nutritional supplement |
| NCT03316157 | Prosure | Nutritional supplement |
| NCT04131426 | Remune | Nutritional supplement |
| NCT04643613 | Protison cancer<br>nutritional formula<br>(PCNF) | Nutritional supplement |
| NCT02877966 | Amixea | Nutritional supplement |
| NCT05771207 | Protein | Protein supplement |
| NCT01046383 | IMN1207 | Cysteine-Rich Non-Denatured Whey Protein Isolate |
| NCT01694602 | 3-methylhistidine<br>(D-3MH) | Amino acid derivative |
| NCT01330823 | L-Carnitine | Amino acid derivative |
| NCT06027242, NCT05023499 | glutamine | Amino acid |
| NCT03253029 | Branched Chain Amino<br>Acid | Amino acid |
| NCT04098237 | Pancrelipase | Digestive enzyme |
| NCT03144128, NCT04864431 | Vitamin D | Vitamin |
| NCT05194397 | Nicotinamide Riboside | Vitamin |
| NCT05264038 | OC514 | "Anticachexia agent" mechanism of<br>action as of yet undisclosed |

**Supplemental File 11.** Biomarkers measured in cancer cachexia clinical trials registered on ClinicalTrials.gov.

| Category | Markers |
| --- | --- |
| Inflammation | CRP, TNF-a, TNF-b, IL-1, IL-1B, IL-2, IL-4, IL-5, IL-6, IL-7, IL-8, IL-10, IL-12, IL-13, IL-15, IL-1ra, IFN Gamma, neutrophil/lymphocyte ratio, activin A, Myostatin, MCP1, ICAM-1, bFGF, G-CSF, Eotaxin, CCL11, IP10, CXCL10, PDGF, VEGF, RANKL, OPG, DLL1, Periostin, CCL5, CCL4, MIP1b, CCL2, CCL3, IGF1, IGFBP3, GDF11, GDF15, p38, IKK, p65, ERK1/2, JNK, p-selectin. |
| Metabolism | Blood glucose, Glucose, Proteasome Activity, Serum Albumin, Pre-albumin, Albumin, Total protein, Fibrinogen, Calcium, Sodium, Chloride, Bicarbonate, LDH, Triglycerides, Glycerol, Free fatty acids, Cholesterol, Transferrin, Hb1Ac, Vitamin A, Vitamin E. |
| Hematology | Hemoglobin, Hematocrit, Erythrocytes, Mean Corpuscular Volume, Leukocytes, Lymphocytes, Platelets, Coagulation test, NK cells, T cells. |
| Liver/Kidney Function | Bilirubin, Aspartate Aminotransferase, Alkaline Phosphatase, Creatinine, ALT, AST, Urea, Protein. |
| Urianalysis | Urine Glucose, Ketones, Urine Protein, Urine Hemoglobin, Urobilinogen, Nitrite, Leukocyte Esterase, Hyaline Casts, Urinary 3-methylhistidine. |
| Homones | Leptin, Ghrelin, Insulin, TSH, LH, FSH, E2, DHEA-S, Testosterone, T4, GLP1, NPY. |
| Molecular/genetic | EGFR, ALK, ROS1, BRAF, HER2, K-RAS, UBE2B, UBE2D2, MaFbx, MuRF1, Nedd4, Mdm2, 26S proteasome subunits, beclin, LAMP, PARP, K48, K63, NRF2, KEAP1, HSP, ER-stress, Pax3, Pax7, Myf5, M-cadherin, myogenin, desmin, SMAD3. |
| Microbiome | Stool sample microbiome analysis, Fecal microbiota, Gut permeability markers, Microbial metabolites. |
| Muscle metabolism/structure | Muscle biopsies, Muscle protein synthesis, ATP concentration in myotubes, Mitochondrial respiration. |
| Bone metabolism/structure | RANKL, Osteoprotegerin, Vitamin D, Calcium, Phosphate, Vitamin K, vitamin K-PT. |
| Biopsies | Adipose tissue biopsy, Muscle biopsy, Tumour biopsy, Subcutaneous adipose tissue, Histological analysis. |
| Others | Total RNA, Reactive oxygen species, Apoptosis markers, Caspase-3, Phosphorylated Akt, Nitrosylated HNE, ATG. |

**Supplemental File 12.** Outcome measures of cachexia in the clinical trials registered on ClinicalTrials.gov by status: Body weight, body composition, physical function, quality of life, nutrition/appetite, molecular, safety and tolerability, and survival.

| Measures of Cachexia |  | Completed<br>with Results<br>(n=52) | Completed<br>Without<br>Results<br>(n=54) | Active<br>(n=62) | Withdrawn<br>(n=23) | Total<br>(n=191) |
| --- | --- | --- | --- | --- | --- | --- |
|  |  | n % | n % | n % | n % | n % |
| Outcome Measures | **Body weight** | 26 (50.0) | 20 (37.0) | 24 (38.7) | 11 (47.8) | 81 (42.4) |
|  | **Body composition** | 34 (65.4) | 26 (48.1) | 45 (72.6) | 14 (60.9) | 119 (62.3) |
|  | Muscle mass | 8 (15.4) | 11 (20.4) | 29 (46.8) | 1 (4.3) | 49 (25.7) |
|  | Lean body mass | 19 (36.5) | 10 (18.5) | 10 (16.1) | 6 (26.1) | 45 (23.6) |
|  | Fat mass | 7 (13.5) | 7 (13.0) | 7 (11.3) | 0 (0.0) | 21 (11.0) |
|  | **Physical function** | 31 (59.6) | 22 (40.7) | 30 (48.4) | 13 (56.5) | 96 (50.3) |
|  | Muscle function | 22 (42.3) | 9 (16.7) | 15 (24.2) | 10 (43.5) | 56 (29.3) |
|  | Physical activity levels | 8 (15.4) | 4 (7.4) | 13 (21.0) | 1 (4.3) | 26 (13.6) |
|  | Fatigue | 8 (15.4) | 3 (5.6) | 2 (3.2) | 1 (4.3) | 14 (7.3) |
|  | **Quality of life (QoL)** | 23 (44.2) | 10 (18.5) | 25 (40.3) | 13 (56.5) | 71 (37.2) |
|  | **Nutrition**<br>**status/appetite** | 22 (42.3) | 10 (18.5) | 15 (24.2) | 11 (47.8) | 58 (30.4) |
|  | **Molecular** | 19 (36.5) | 21 (38.9) | 38 (61.3) | 9 (39.1) | 87 (45.5) |
|  | **Safety & tolerability** | 13 (25.0) | 11 (20.4) | 10 (16.1) | 7 (30.4) | 41 (21.5) |
|  | **Survival** | 10 (19.2) | 11 (20.4) | 12 (19.4) | 4 (17.4) | 37 (19.4) |

n represents the number of outcomes stated during the clinical trials (body weight, body composition, physical function, quality of life, nutrition status/appetite, molecular, safety and tolerability, survival, or others). % refers to the percentage of clinical trials relative the total of clinical trials within each status (completed with results, completed without results, active, or withdrawn). The % in the “Total” column shows the percentage relative to the total of trials included in the study.

**Supplemental File 13.** Measurement tools used to investigate cachexia by the clinical trials registered on ClinicalTrials.gov by status.

| Measures of Cachexia |  | Completed with Results (n=52) | Completed Without Results (n=54) | Active (n=62) | Withdrawn (n=23) | Total (n=191) |
| --- | --- | --- | --- | --- | --- | --- |
|  |  | n % | n % | n % | n % | n % |
| Measurement Tools | **Body Composition Assessment** |  |  |  |  |  |
|  | CT | 7 (13.5) | 7 (13.0) | 19 (30.6) | 2 (8.7) | 35 (18.3) |
|  | BIA | 12 (23.1) | 8 (14.8) | 8 (12.9) | 5 (21.7) | 33 (17.3) |
|  | DXA | 8 (15.4) | 3 (5.6) | 8 (12.9) | 5 (21.7) | 24 (12.6) |
|  | MRI | 2 (3.8) | 2 (3.7) | 4 (6.5) | 1 (4.3) | 9 (4.7) |
|  | Skinfold | 4 (7.7) | 0 (0.0) | 1 (1.6) | 3 (13.0) | 8 (4.2) |
|  | MUAC | 3 (5.8) | 1 (1.9) | 0 (0.0) | 1 (4.3) | 5 (2.6) |
|  | PET | 0 (0.0) | 1 (1.9) | 3 (4.8) | 0 (0.0) | 4 (2.1) |
|  | Waist-Hip ratio | 0 (0.0) | 1 (1.9) | 2 (3.2) | 0 (0.0) | 3 (1.6) |
|  | **Physical Function Assessment** |  |  |  |  |  |
|  | Hand Grip Strenght | 19 (36.5) | 7 (13.0) | 15 (24.2) | 9 (39.1) | 50 (26.2) |
|  | Walk test | 5 (9.6) | 6 (14.8) | 13 (21.0) | 1 (4.3) | 25 (13.1) |
|  | Sit-to-stand | 3 (5.8) | 3 (5.6) | 9 (14.5) | 3 (13.0) | 18 (9.4) |
|  | Stair Climb | 5 (9.6) | 2 (3.7) | 3 (4.8) | 2 (8.7) | 12 (6.3) |
|  | SPPB | 0 (0.0) | 5 (9.3) | 3 (4.8) | 0 (0.0) | 8 (4.2) |
|  | Accelorometry | 3 (5.8) | 0 (0.0) | 1 (1.6) | 0 (0.0) | 4 (2.1) |
|  | **Symptom and QoL Assessment** |  |  |  |  |  |
|  | ECOG | 23 (44.2) | 12 (22.2) | 19 (30.6) | 9 (39.1) | 63 (33.0) |
|  | EORTC QLQ | 12 (23.1) | 8 (14.8) | 13 (21.0) | 7 (30.4) | 40 (20.9) |
|  | FAACT | 11 (21.2) | 7 (13.0) | 7 (11.3) | 1 (4.3) | 26 (13.6) |
|  | Karnofsky scale | 5 (9.6) | 3 (5.6) | 4 (6.5) | 5 (21.7) | 17 (8.9) |
|  | FACIT-F | 4 (7.7) | 2 (3.7) | 2 (3.2) | 0 (0.0) | 8 (4.2) |
|  | **Nutrition and Appetite Assessment** |  |  |  |  |  |
|  | Dietary recall/food diary | 4 (7.7) | 5 (9.3) | 5 (8.1) | 4 (17.4) | 18 (9.4) |
|  | PG- SGA | 2 (3.8) | 1 (1.9) | 2 (3.2) | 2 (8.7) | 7 (3.7) |
|  | VAS | 4 (7.7) | 0 (0.0) | 0 (0.0) | 2 (8.7) | 6 (3.1) |
|  | SNAQ | 2 (3.8) | 1 (1.9) | 1 (1.6) | 0 (0.0) | 4 (2.1) |
|  | **Molecular Assessment** |  |  |  |  |  |
|  | Blood sample | 17 (32.7) | 25 (46.3) | 34 (54.8) | 9 (39.1) | 85 (44.5) |
|  | Muscle biopsy | 0 (0.0) | 7 (13.0) | 11 (17.7) | 3 (13.0) | 21 (11.0) |

CT: Computed Tomography, BIA: Bioelectrical Impedance Analysis, DXA: Dual Energy X-ray Absorptiometry, MRI: Magnetic Resonance Imaging, MUAC: Mid-Upper Arm Circumference, PET: Positron Emission Tomography, SPPB: Short Physical Performance Battery, ECOG: Eastern Cooperative Oncology Group, EORTC QLQ: Research and Treatment of Cancer Quality of Life (QoL) Questionary, FAACT: Functional Assessment of Anorexia/Cachexia Treatment, FACIT-F: Functional Assessment of Chronic Illness Therapy – Fatigue, PG- SGA: Patient-Generated Subjective Global Assessment, VAS: Visual Analog Scale, SNAQ: Simplified Nutritional Appetite Questionnaire.

